# No evidence of association between schools and SARS-CoV-2 second wave in Italy

**DOI:** 10.1101/2020.12.16.20248134

**Authors:** Sara Gandini, Maurizio Rainisio, Maria Luisa Iannuzzo, Federica Bellerba, Francesco Cecconi, Luca Scorrano

## Abstract

**Background:** During the Covid19 pandemic, school closure has been mandated in analogy to its known effect against influenza, but it is unclear whether schools are early amplifiers of Covid19 cases.

**Methods:** We performed a cross-sectional and prospective cohort study in Italy. We used databases from the Italian Ministry of Education containing the number of new positive SARS-CoV-2 cases per school from September 20 to November 8, 2020 to calculate incidence among students and staff. We calculated incidence across each age group using databases from the Veneto Region system of SARS-CoV-2 cases notification in the period August 26- November 24, 2020. We used a database from the Veneto Region system of SARS-CoV-2 secondary cases tracing in Verona province schools to estimate number of tests, the frequency of secondary infections at school by type of index case and the ratio positive cases/ number of tests per school institute using an adjusted multivariable generalized linear regression model. We estimated the reproduction number R_t_ at the regional level from the Italian Civil Protection of regional SARS-CoV-2 cases notification database in the period 6 August-2 December 2020.

**Findings:** From September 12 to November 7 2020, SARS-CoV-2 incidence among students was lower than that in the general population of all but two Italian regions. Secondary infections were <1%, and clusters of >2 secondary cases per school were 5-7% in a representative November week. Incidence among teachers was greater than in the general population. However, when compared with incidence among similar age groups, the difference was not significant (P=0.23). Secondary infections among teachers were more frequent when the index case was a teacher than a student (38% vs. 11%, P=0.007). From August 28 to October 25 in Veneto where school reopened on September 14, the growth of SARS-CoV- 2 incidence was lower in school age individuals, maximal in 20-29 and 45-49 years old individuals. The delay between the different school opening dates in the different Italian regions and the increase in the regional Covid19 reproduction number R_t_ was not uniform. Reciprocally, school closures in two regions where they were implemented before other measures did not affect the rate of R_t_ decline.

**Interpretation:** Our analysis does not support a role for school opening as a driver of the second wave of SARS-CoV-2 epidemics in Italy, a large European country with high SARS-CoV-2 incidence.

**Research in context:** *Evidence before this study:* The role of schools and at large of children as amplifiers of the Covid19 pandemics is debated. Despite biological and epidemiological evidence that children play a marginal role in Sars-CoV-2 spread, policies of school closures have been predicated, mostly based on the temporal coincidence between school reopening in certain countries and Covid19 outbreaks. Whether schools contributed to the so called “second wave” of Covid19 is uncertain. Italy’s regionalized calendar of school reopening and databases of positivity at school allows to estimate the impact of schools on the increase of Sars-CoV-2 that occurred in autumn 2020.

*Added value of this study:* We found that incidence among students is lower than in the general population and that whereas incidence among teachers appears higher than that in the general population, it is comparable to that among individuals of the same age bracket. Moreover, secondary infections at school are rare and clusters even less common. The index case of a secondary teacher case is more frequently a teacher than a student. In Veneto, during the first phase of the second wave incidence among school age individuals was low as opposed to the sustained incidence among individuals of 45-49 years. Finally, the time lag between school opening and Rt increase was not uniform across different Italian regions with different school opening dates, with lag times shorter in regions where schools opened later.

*Implications of the available evidence:* These findings contribute to indicate that Covid19 infections rarely occur at school and that transmission from students to teachers is very rare. Moreover, they fail to support a role for school age individuals and school openings as a driver of the Covid19 second wave. Overall, our findings could help inform policy initiatives of school openings during the current Covid19 pandemic.

## Introduction

School closures represent a widespread nonpharmacological intervention (NPI) in the context of the current Coronavirus Disease 2019 (Covid19) pandemic. In Italy, schools have been closed for more than half of the 2019-2020 school year and during the second Covid19 wave high schools have been closed again, with students switching to “integrated digital learning” nationwide since November 6, 2020. The rationale for such a NPI has mostly been drawn from the reported beneficial effect of school closure during influenza pandemics ^1^, even if the debate was still open ^2^. However, while children’s immune system is naïve to influenza antigens, making them a known reservoir of influenza infection, they do not appear as affected by Covid19 as adults, representing a small fraction of documented Covid19 cases. Similarly to SARS-CoV and MERS-CoV, SARS-CoV-2 affects less children, where symptoms are fewer, disease less severe and case-fatality rates much lower ^3,45^.

Several biological factors might contribute to the reduced Covid19 risk of children: first, children express significantly fewer ACE2 receptors – the entry point of SARS-CoV-2 into human cells – compared to adults ^6^; second, they are commonly exposed to other seasonal coronaviruses and develop both humoral and cellular cross-immunity ^7^. Children appear therefore less susceptible to the infection, and when infected have a preformed arsenal of neutralizing cross reactive antibodies that might reduce the likelihood of transmitting the virus. This biological evidence is mirrored in several epidemiological studies. A meta-analysis of 32 studies from different countries suggests that children have a lower susceptibility to SARS-CoV-2 infection compared with adults ^8^. An age-structured mathematical model applied to epidemic data from China, Italy, Japan, Singapore, Canada and South Korea estimates that individuals younger than 20 years of age display half the chance of being infected than adults ^9^. In the context of households (the most common route of secondary infection) chances of transmission from children to adults are low and the spread seldom starts from children. In a large study including 15,771 children aged 1 to 18 years living in Germany, almost two-thirds of children living with virus-positive family members were negative for SARS-CoV-2 antibodies and virus tests, suggesting that transmission to children is infrequent ^10^. In only three families (9.7%) among 31 household transmission cases that involved children in China, Singapore, USA, Vietnam, and South Korea the child represented the index case ^11^. In a meta-analysis of all contact-tracing studies up to 16^th^ May 2020, children are less susceptible to SARS-CoV-2 than adults, with 56% lower odds of being an infected contact [Pooled OR=0.44 (95%CI 0.29, 0.69)] ^8^. In the Italian town of Vo’ Euganeo, where 70% of the population was screened and 2.6% of the population resulted positive, no child below 10 years of age was found positive, even if these children lived in the same household with a positive individual ^12^. In a large cohort study on 12 million people in the UK, the risk of infecting and becoming infected with SARS-COV-2 grew with age. In the same study, risk of contracting the virus was not higher than that of the rest of the population for >9 million adults younger than 65 living with children up to 11 years of age. The risk increased slightly for those who lived with adolescents aged 12 to 18, but this risk did not correspond to a greater lethality in case of SARS-CoV-2 infection. Indeed, there was no significant effect of the school closure on the trend of the epidemic in the families analyzed compared to the rest of the population ^13^.

Despite evidence supporting a marginal role for children in Covid19 pandemic, school openings (or re-openings) have been considered an issue. This notion has been based on modeling studies and systematic reviews showing conflicting results on whether school closures were efficacious in curtailing the general incidence of infection ^14,15^. Moreover, adolescents were reported to be just as likely as adults to spread the virus ^16^, and in one study levels of Sars-CoV-2 genetic material in the upper respiratory tract of children below 5 years with mild to moderate Covid19 was more than in children 6-17 and adults ^17^. Furthermore, in a Covid19 outbreak at a summer camp in Georgia, children of all ages were found to be highly susceptible to infection: 51% of the campers ages 6 to 10 tested positive, as did 44% of those ages 11 to 17 ^18^. In Israel, schools fully reopened on May 17, 2020 and ten days later, a major outbreak of coronavirus disease (Covid19) occurred in a high school; temporal correlation between school openings and the second wave was interpreted as a causal link ^19^. By extension, policymakers (as well as the lay public) attribute to school openings a key role in amplifying infection rates in the general population. This opinion is particularly widespread in Italy, where schools remained closed from February 25 in Northern Italian regions (from March 9 nationwide) until September, when they reopened in different days across different regions. Despite the implementation of a very strict infection risk mitigation protocol, school openings have been regarded as a key factor in the amplification of the second Covid19 wave and their closure has been predicated by several data analysts. Consequently, high schools nationwide and, in certain regions, the second and third year of middle schools (the Italian middle school curriculum lasts 3 years) have been closed since November 6. In other regions (Campania and Apulia), closure of all schools including elementary and kindergarten has been mandated since October 16 and 30, respectively. In Lombardy, high schools have been closed since October 26. However, whether school openings played a crucial role in the second wave of Covid19 infections remains to be ascertained. Italy was in a privileged position to investigate this possibility: school calendars are regional and starting dates differ among different regions by up to 17 days.

The aims of this study were to investigate the overall incidence of SARS-CoV-2 infection among students and teachers, as well as whether there was an association between the increase in transmissibility of SARS-CoV-2 (measured as reproduction number R_t_) and dates of school openings in different Italian Regions. We also estimated the incidence of SARS-CoV-2 by age in Veneto and the incidence of SARS-CoV-2 positive students, teachers, and other staff in public and private schools in two weeks between the end of November and beginning of December in the Italian regions. We calculated the rate of secondary infections per number of tests and frequency of clusters identified during contact tracing activity in a large sample of Italian Schools. We also estimated the frequency of secondary infections in teachers by type of index case (student, teacher or staff).

## Methods

### Study design

This was a cross-sectional and prospective cohort study. Data collected within the comprehensive, national reporting system put in place by the Italian Ministry of Education (Ministero dell’Istruzione - MI) to gather information from school Principals every week for each comprehensive private and state institute. All institutes are required to inform the Department of Prevention of the local unit (AULSS) of the National Health System responsible for tracing and the MI when they suspect or identify a case or outbreak of Covid19. AULSS then perform a risk assessment and decide on any additional investigation and infection control measure, based on factors such as the number of new positive subjects, disease severity, and potential transmission at school. AULSS record each event within each region in an online national database of public health management. MI and AULSS have legal permission to process patient confidential information for national surveillance of communicable diseases.

For outbreaks, direction of transmission from the index case to secondary cases was inferred based on the date of symptom onset for symptomatic individuals and date of testing for asymptomatic individuals. We evaluated associations between event measures in educational settings, regional Covid19 incidence, and other regional characteristics to identify possible predictors for cases and outbreaks.

We used a database with data collected by MI containing the number of new positive SARS-CoV-2 cases per school from September 20 to November 8, 2020. This database reports the incidence in the first cycle (kindergarten, elementary and middle school) and second cycle (high school) by region. Data was retrieved from 7,976 public school institutes (97% of total), accounting for 7,376,698 students, 775,451 teachers and 206,120 staff members.

We also analyzed data of SARS-CoV-2 incidence in schools in the period 23-28 November, 2020 in a sample of 6,827 public institutes (81.6% of the total) and 7,035 private institutes (55.6% of the total institutes). Furthermore, we analyzed data collected by MI from contact tracing in the monitored schools (from 23 November to 5 December 2020). Information were retrieved from 5.971 (45%) institutes in the first week, and 7,035 (55.6%) institutes in the second week (public and private institutes), accounting for 423.516 and 496,289 students in the first and second week, respectively.

We calculated regional Covid19 incidence using national SARS-CoV-2 RT-PCR data collated by AULSS for Covid19 surveillance, and regional population estimates from the Office for National Statistics (Istituto Nazionale di Statistica, ISTAT).

SARS-CoV-2 positive incidence rates were calculated for staff and students attending an educational setting, irrespective of whether the infection was acquired within or outside the educational setting. attendance denominators for educational settings were obtained from the MI open database (https://dati.istruzione.it/opendata/). For event rate and incidence rates calculations, denominators were drawn from MI enrollment figures.

### Statistical methods

Mean, median values, inter-quartile ranges, standard deviations and box-plots for continuous variables and absolute and relative frequencies for categorical variables are presented. Differences among groups for continuous variables were tested by means of the non-parametric Wilcoxon-rank sum test and differences for categorical variables were tested by means of the Chi-square test.

Incidence rates of secondary infections were defined as number of cases/number of tests occurring the same week after a SARS-COV-2 positive student or teacher was found. LSmeans, 95%CI and P-values of rate of secondary infections and number of positive test per institute and week are estimated with multivariable generalized linear regression model adjusted for week of test and density of the region, weighted for the number of tests released in each institute to trace close contacts. Square root transformations were carried out to achieve normality of residuals of full models.

Incidence rates were calculated as the sum of all new positives in each week, divided by the size of the population. We work out the cases per 10,000 (a standard epidemiological way of presenting incidence) by dividing the number of cases by the population in each age group (estimates are from ISTAT, 2018).

To generate the incidence heatmap, a matrix of the weekly incidence referred to individual age ranges was calculated. Using Excel, individual cells were color coded in a 3 color scale (green-beige-red) of increasing weekly incidence rate. To generate the heatmap of distance between age brackets, the same matrix was fed to the Heatmapper algorithm (www.heatmapper.ca) and we selected to calculate the distance between rows and columns using the Euclidean Distance Measurement Method.

Transmissibility was measured by the reproduction number R_t_, as the average number of secondary cases caused by an infected individual. We estimated R_t_ over the months incorporating uncertainty in the distribution of the serial interval (the time between the onset of symptoms in a primary case and the onset of symptoms in secondary cases)^20^. R_t_ was computed using EpiEstim ^20^ with parameters from the first wave in Italy as defined by Merler and coworkers ^21^ (serial interval: 6.6, gamma: 4.9). R_t_ was computed using the number of new cases/day in each region (physician referral and screening due to local campaigns and to contact tracing). Data were retrieved from the database of the Italian Civil Protection (https://github.com/pcm-dpc/Covid19). In all graphs, R_t_ values are reported as median values for a 7-day posterior moment with 95% credible intervals. When an NPI was introduced and school opening occurred, their effect on R_t_ was referred to the first day of the corresponding 7-day moment. For example, if schools opened on September 14, their effect on R_t_ was introduced from the period September 14-20.

Since previous analyses suffer of ecological bias, we also performed a prospective study on datasets extracted from the Veneto Region system of SARS-CoV-2 cases notification. We stratified incidence of newly reported Covid19 cases for age from late August to late October 2020, when overall Covid19 incidence in Veneto increased from ∼2/10,000 to ∼35/10,000. We first divided the population into infants (0-4 years of age), students (5-19), young adults (20-34), adults (35-59), seniors (60-74) and elderly (75+) and calculated daily incidence of newly reported cases in these age categories. We also extracted information regarding secondary infections among teachers in 339 schools in Verona and province from November 25 to December 21, by type of index case. Statistical analyses were performed with Statistical Analysis System Version 9.4 (SAS Institute, Cary, NC, USA) or with OriginPro 2021 (OriginLab, Northampton, MA, USA).

## Results

### School demographics and the 2020-21 school year in Italy

According to the National Statistical Institute (ISTAT), 9,150,518 students attended the different school cycles in 2019 in Italy. These cycles include kindergarten (*scuola dell’infanzia*, attended by 3-5 years old children), elementary (*scuola primaria* attended by 6-10 years old children), middle (*scuola secondaria di primo grado* attended by 11-13 years old children) and high school (*scuola secondaria di secondo grado* attended by 14-18 years old children). Education is compulsory from 6 to 16 years of age. Pre-elementary school education that includes kindergarten as well as nurseries (*asili nido*, attended by children 0- 2 years old) is not compulsory. On average, students represented 15% of the population of each of the 20 Italian regions and two autonomous Provinces (range: 10.7%-19%; Table 1). While kindergartens and nurseries started nationwide on September 1^st^, the calendarized opening day of all other schools differed among regions. In most regions, schools started on September 14; in a second group of regions schools opened on September 24; in two other regions on September 16 or 22 (Table 2). The Italian Government mandated a protocol to minimize risk of Covid19 diffusion that followed most of the strictest recommendations ^22^. Measures included temperature control and hand hygiene at the school entrance; unidirectional flows of students; mask mandate for all personnel and students in common areas and for high school students also when seated at their desks (and always for teachers, combined with face-shields in certain settings), compulsory 1m seat to seat distance, frequent classroom natural ventilation, ban on school sports and music, reduced duration of school hours and reduced school duration. Moreover, Covid19 positive staff members must promptly inform the school Principal. Similarly, parents must promptly report any case of Covid19 positivity in their children to the schools and Principals must coordinate with local units of the National Health System to perform secondary screenings among staff/students, or to mandate quarantine for 10 days with a swab to all quarantined students/personnel before re-admitting them to the school premises. Notwithstanding these rules, school opening has been accounted as the driver of the second Covid19 wave by the popular press as well as by opinion makers, calling for a careful analysis of Covid19 diffusion and contact tracing among children in schools, as well as of the association between dates of school openings and qualitative changes in the R_t_ trend direction (1^st^ derivative of R_t_), a widely accepted proxy of epidemic driver.

**Table 1.** Demographics of Italian Regions and autonomous Provinces. Data are from the National Statistical Institute (ISTAT). In Italy, elementary school starts at 6, middle school at 11, high school at 14 years of age.

| Region | Popula<br>tion | Preschool<br>students<br>(%) |  | Elementary<br>school<br>students<br>(%) |  | Middle<br>school<br>students<br>(%) |  | High<br>school<br>students<br>(%) |  | Studen<br>ts/<br>popula<br>tion<br>(%) |
| --- | --- | --- | --- | --- | --- | --- | --- | --- | --- | --- |
| **Abruzzo** | 1,305,770 | 48,397 | 3.7 % | 55,893 | 4.3 % | 34,881 | 2.7 % | 58,308 | 4.5 % | 15.1% |
| **Apulia** | 4,008,296 | 181,674 | 4.5 % | 178,761 | 4.5 % | 115,152 | 2.9 % | 205,348 | 5.1 % | 17.0% |
| **Basilicata** | 556,934 | 19,710 | 3.5 % | 14,110 | 2.5 % | 14,696 | 2.6 % | 26,640 | 4.8 % | 13.5% |
| **Bolzano** | 532,080 | 27,742 | 5.2 % | 27,592 | 5.2 % | 17,097 | 3.2 % | 28,846 | 5.4 % | 19.0% |
| **Calabria** | 1,924,701 | 80,534 | 4.2 % | 85,450 | 4.4 % | 54,642 | 2.8 % | 77,850 | 4.0 % | 15.5% |
| **Campania** | 5,785,861 | 254,097 | 4.4 % | 232,042 | 4.0 % | 183,729 | 3.2 % | 324,049 | 5.6 % | 17.2% |
| **Emilia-Romagna** | 4,467,118 | 207,566 | 4.6 % | 203,083 | 4.5 % | 87,735 | 2.0 % | 200,680 | 4.5 % | 15.6% |
| **Friuli-Venezia Giulia** | 1,211,357 | 34,169 | 2.8 % | 50,546 | 4.2 % | 22,584 | 1.9 % | 43,230 | 3.6 % | 12.4% |
| **Lazio** | 5,865,544 | 239,656 | 4.1 % | 223,071 | 3.8 % | 168,949 | 2.9 % | 270,075 | 4.6 % | 15.4% |
| **Liguria** | 1,543,127 | 59,214 | 3.8 % | 48,338 | 3.1 % | 38,327 | 2.5 % | 64,141 | 4.2 % | 13.6% |
| **Lombardy** | 10,103,969 | 256,204 | 2.5 % | 475,220 | 4.7 % | 208,087 | 2.1 % | 477,029 | 4.7 % | 14.0% |
| **Marche** | 1,518,400 | 66,271 | 4.4 % | 66,740 | 4.4 % | 29,095 | 1.9 % | 68,507 | 4.5 % | 15.2% |
| **Molise** | 302,265 | 12,214 | 4.0 % | 11,544 | 3.8 % | 7,484 | 2.5 % | 10,903 | 3.6 % | 13.9% |
| **Piedmont** | 4,341,375 | 137,009 | 3.2 % | 151,981 | 3.5 % | 117,142 | 2.7 % | 156,974 | 3.6 % | 13.0% |
| **Sardinia** | 1,630,474 | 51,318 | 3.1 % | 63,957 | 3.9 % | 40,501 | 2.5 % | 19,189 | 1.2 % | 10.7% |
| **Sicily** | 4,968,410 | 247,970 | 5.0 % | 190,547 | 3.8 % | 147,430 | 3.0 % | 259,111 | 5.2 % | 17.0% |
| **Tuscany** | 3,722,729 | 135,146 | 3.6 % | 130,853 | 3.5 % | 101,638 | 2.7 % | 135,178 | 3.6 % | 13.5% |
| **Trento** | 542,739 | 19,206 | 3.5 % | 26,771 | 4.9 % | 16,483 | 3.0 % | 27,833 | 5.1 % | 16.6% |
| **Umbria** | 880,285 | 37,363 | 4.2 % | 31,048 | 3.5 % | 24,520 | 2.8 % | 39,075 | 4.4 % | 15.0% |
| **Valle D'Aosta** | 125,501 | 4,647 | 3.7 % | 5,740 | 4.6 % | 3,662 | 2.9 % | 4,758 | 3.8 % | 15.0% |
| **Veneto** | 4,907,704 | 225,722 | 4.6 % | 223,780 | 4.6 % | 142,348 | 2.9 % | 233,716 | 4.8 % | 16.8% |
| **Italy** | 60,244,639 | 2,345,829 | 4% | 2,497,067 | 4% | 1,576,182 | 3% | 2,731,440 | 5% | 15% |

**Table 2:** Dates of School opening in the 21 Italian Regions and autonomous Provinces (Trento and Bolzano).

| School Opening | Sept. 7 | Sept. 14 | Sept. 16 | Sept. 22 | Sept. 24 |
| --- | --- | --- | --- | --- | --- |
| Region/Autonomous Province | Bolzano | Emilia-Romagna | Friuli Venezia Giulia | Sardinia | Abruzzo |
|  |  | Lazio |  |  | Apulia |
|  |  | Liguria |  |  | Basilicata |
|  |  | Lombardy |  |  | Calabria |
|  |  | Marche |  |  | Campania |
|  |  | Molise |  |  |  |
|  |  | Piedmont |  |  |  |
|  |  | Sicily |  |  |  |
|  |  | Tuscany |  |  |  |
|  |  | Trento |  |  |  |
|  |  | Umbria |  |  |  |
|  |  | Valle D'Aosta |  |  |  |
|  |  | Veneto |  |  |  |

### Incidence of Covid19 among students is lower than in the general population

To first gain insight into the diffusion of Covid19 in Italian Schools, we analyzed a database from the Italian Ministry of Education (MI). We used this database to calculate the incidence of new positives in the period and per week among students, teachers, and non-teaching staff members of elementary, middle and high schools. We compared these data to the incidence of SARS-CoV-2 positivity in the general population for each region. The incidence of positives among students was lower than that in the population (overall incidence: 108/10,000), irrespective of whether we analyzed elementary and middle schools (incidence: 66/10,000), or high schools (incidence: 98/10,000). Incidence of new positives among elementary and middle school students was on average 38.9% lower than in the general population in all Italian regions but Lazio (Figure 1A). In the case of high schools, incidence of new positives among the students was 9% lower to that of the general population (Figure 1B). In the three regions of Lazio, Marche, and Emilia-Romagna, it was higher than in the general population. Among teachers and non-teaching staff incidence was 2-fold higher than that observed in the general population (approx. 220/10,000, Figure 1C). These data indicate that students are largely protected from SARS-CoV-2 infection, irrespective of their school cycle. Conversely, infection appears to be more widespread among teachers and staff members of schools than in the general population. It shall be noted that while teachers share classrooms for several hours with students, staff members include administrative personnel and janitors who seldom interact with students.

**Figure 1.**
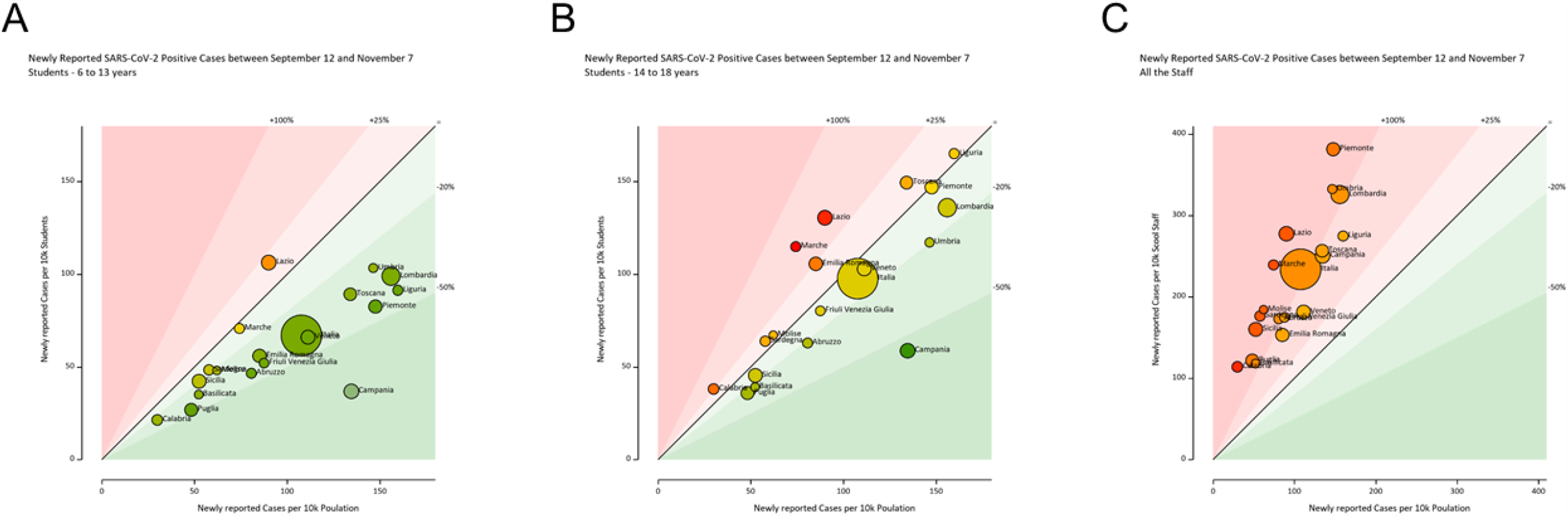
Incidence of SARS-CoV-2 positives is lower among schoolers than in the general population. Bubble graphs of SARS-CoV-2 incidence between September 12 and November 7 for student in elementary schools (A), middle schools (B) and staff members (teaching and non-teaching, C) in Italian regions and autonomous provinces compared to the incidence in the general population. Size of bubbles is proportional to the measured incidence in the analyzed school populations. The 45° line indicates equivalence between general population and school population incidence. Bubbles are color-coded in a green-yellow-red gradient of analyzed population/general population ratio.

We next used a second database in which the MI collected the number of new cases in the period 23-28 November in a sample of 6,827 public (81.6% of the total) and 7,035 private institutes (55.6% of the total). This database offers a snapshot of the distribution of new cases in a limited timeframe during the peak of the second Covid19 wave. New positive subjects were found mostly among teachers and staff members: SARS-CoV-2 positives were 0.32% of students, 1.52% of teachers and 1.96% of staff members (Table 3, Figure S1). The highest rate was found in Molise and the lowest in Calabria. Incidences of new cases in kindergarten were 0.21% in pupils and 2.35% among teachers (P<0.001), in elementary schools were 0.35% among children and 1.83% among teachers (P<0.001). In middle schools, 0.45% students and 1.60% teachers were found positive (P<0.001, Table 4 and 5). Similar incidence rates were found in private schools (Table S3-S4), except for a slightly lower rate among staff members (1.67%, Table S1). This database allowed us also to investigate how often the communication of a positive case elicited quarantine for students/staff members. A quarantine period was requested for 1.92% of students, 2.30% of teachers and 2.56% of staff members of the analyzed public schools (Table 6). In private schools, rates of quarantines were very similar, except for a slightly higher rate for children (2.65%, Table S4). These data indicate that even during the peak of the second Covid19 wave, students were less infected than adults in school establishments, and that overall, the quarantine system was widespread, vis-à-vis a very low rate of positivity among students.

**Table 3.** Incidence in students, teachers, and other school personnel by region, from kindergarten to middle school in the period 23-28 November. Trentino-Alto Adige e Valle d’Aosta do not use the MI national system.

| REGION | New positives among students | % of positive students | New positives among teachers | % of positive teachers | New positives among other school personnel | % of positives among other school personnel |
| --- | --- | --- | --- | --- | --- | --- |
| Abruzzo | 413 | 0.32% | 680 | 1.35% | 224 | 1.81% |
| Apulia | 778 | 0.17% | 817 | 1.13% | 35 | 1.17% |
| Basilicata | 267 | 0.46% | 657 | 1.68% | 394 | 2.25% |
| Calabria | 252 | 0.12% | 209 | 0.80% | 72 | 0.95% |
| Campania | 1,004 | 0.16% | 212 | 1.05% | 20 | 0.98% |
| Emilia Romagna | 1,990 | 0.47% | 2,004 | 2.03% | 594 | 2.53% |
| Friuli Venezia Giulia | 381 | 0.38% | 749 | 1.53% | 24 | 2.09% |
| Lazio | 1,579 | 0.31% | 832 | 1.29% | 258 | 1.58% |
| Liguria | 430 | 0.32% | 201 | 1.36% | 78 | 1.84% |
| Lombardy | 3,791 | 0.40% | 243 | 1.57% | 196 | 2.16% |
| Marche | 509 | 0.30% | 246 | 1.27% | 171 | 1.48% |
| Molise | 157 | 0.49% | 1,195 | 2.61% | 335 | 2.78% |
| Piedmont | 1,215 | 0.30% | 752 | 1.28% | 79 | 1.52% |
| Sardinia | 429 | 0.28% | 143 | 1.24% | 52 | 1.38% |
| Sicily | 1,887 | 0.35% | 625 | 1.47% | 258 | 2.06% |
| Tuscany | 1,117 | 0.32% | 59 | 1.43% | 261 | 2.05% |
| Umbria | 286 | 0.28% | 89 | 1.22% | 72 | 1.36% |
| Veneto | 2,068 | 0.44% | 182 | 1.67% | 68 | 2.19% |
| **TOTAL** | **18,553** | **0.32%** | **9,895** | **1.52%** | **3,191** | **1.96%** |

**Table 4.** Incidence in students and teachers by region in kindergarten and elementary school in the period 23-28 November. Trentino-Alto Adige e Valle d’Aosta do not use the MI national system

|  | Kindergarten |  |  |  | Elementary schools |  |  |  |
| --- | --- | --- | --- | --- | --- | --- | --- | --- |
| REGION | New positives among students | % of positive students | New positives among teachers | % of positive teachers | New positives among students | % of positive students | New positives among teachers | % of positive teachers |
| Abruzzo | 42 | 0.20% | 54 | 2.40% | 202 | 0.50% | 78 | 1.70% |
| Apulia | 102 | 0.16% | 146 | 2.13% | 209 | 0.15% | 248 | 1.67% |
| Basilicata | 10 | 0.12% | 12 | 1.32% | 98 | 0.61% | 29 | 1.38% |
| Calabria | 42 | 0.14% | 44 | 1.22% | 99 | 0.16% | 75 | 0.95% |
| Campania | 49 | 0.06% | 152 | 1.46% | 168 | 0.09% | 294 | 1.38% |
| Emilia Romagna | 103 | 0.27% | 72 | 1.80% | 677 | 0.49% | 255 | 1.64% |
| Friuli Venezia Giulia | 26 | 0.25% | 31 | 2.52% | 117 | 0.36% | 82 | 2.17% |
| Lazio | 119 | 0.21% | 149 | 2.35% | 525 | 0.32% | 344 | 1.68% |
| Liguria | 21 | 0.14% | 39 | 2.35% | 137 | 0.33% | 85 | 1.67% |
| Lombardy | 240 | 0.28% | 368 | 3.89% | 1,598 | 0.47% | 915 | 2.39% |
| Marche | 34 | 0.14% | 48 | 1.74% | 129 | 0.24% | 72 | 1.17% |
| Molise | 11 | 0.23% | 7 | 1.22% | 68 | 0.69% | 25 | 2.02% |
| Piedmont | 162 | 0.33% | 249 | 4.26% | 396 | 0.29% | 445 | 2.68% |
| Sardinia | 37 | 0.21% | 32 | 1.40% | 174 | 0.39% | 87 | 1.44% |
| Sicily | 155 | 0.20% | 144 | 1.74% | 697 | 0.41% | 323 | 1.58% |
| Tuscany | 118 | 0.27% | 126 | 2.63% | 414 | 0.39% | 254 | 2.00% |
| Umbria | 54 | 0.39% | 28 | 1.92% | 131 | 0.41% | 65 | 1.74% |
| Veneto | 117 | 0.38% | 88 | 2.50% | 747 | 0.47% | 308 | 1.79% |
| **TOTALE** | **1,442** | **0.21%** | **1,789** | **2.35%** | **6,586** | **0.35%** | **3,984** | **1.83%** |

**Table 5.** Incidence in students and teachers by region in middle school in the period 23-28 November. Trentino-Alto Adige e Valle d’Aosta do not use the MI national system

| REGION | New positives among students | % of positive students | New positives among teachers | % of positive teachers |
| --- | --- | --- | --- | --- |
| Abruzzo | 110 | 0.43% | 70 | 2.17% |
| Apulia | 175 | 0.17% | 136 | 1.22% |
| Basilicata | 117 | 1.03% | 31 | 2.09% |
| Calabria | 81 | 0.18% | 64 | 1.08% |
| Campania | 188 | 0.13% | 168 | 1.00% |
| Emilia Romagna | 679 | 0.70% | 160 | 1.83% |
| Friuli Venezia Giulia | 123 | 0.52% | 39 | 1.67% |
| Lazio | 642 | 0.54% | 150 | 1.13% |
| Liguria | 224 | 0.70% | 48 | 1.43% |
| Lombardy | 1,014 | 0.44% | 454 | 1.92% |
| Marche | 183 | 0.47% | 64 | 1.64% |
| Molise | 55 | 0.78% | 17 | 1.99% |
| Piedmont | 295 | 0.32% | 309 | 2.98% |
| Sardinia | 157 | 0.46% | 65 | 1.46% |
| Sicily | 715 | 0.58% | 212 | 1.36% |
| Tuscany | 327 | 0.44% | 166 | 2.07% |
| Umbria | 59 | 0.27% | 20 | 0.85% |
| Veneto | 775 | 0.71% | 179 | 1.60% |
| **TOTALE** | **5,919** | **0.45%** | **2,352** | **1.60%** |

**Table 6.** Quarantines in students, teachers and other personnel by region, from kindergarten to middle school in the period 23-28 November. Trentino-Alto Adige e Valle d’Aosta do not use the MI national system.

| REGION | Cases of quarantine among students | % of quarantine among students | Cases of quarantine among teachers | % of quarantine among teachers | Cases of quarantine among other personnel | % of quarantine among other personnel |
| --- | --- | --- | --- | --- | --- | --- |
| Abruzzo | 2,532 | 1.95% | 308 | 2.68% | 55 | 2.71% |
| Apulia | 7,574 | 1.64% | 816 | 1.92% | 521 | 2.22% |
| Basilicata | 455 | 0.79% | 107 | 1.47% | 164 | 2.17% |
| Calabria | 1,556 | 0.74% | 374 | 1.43% | 64 | 1.70% |
| Campania | 1,099 | 0.18% | 620 | 0.86% | 80 | 1.54% |
| Emilia Romagna | 9,969 | 2.36% | 1,231 | 2.69% | 247 | 2.72% |
| Friuli Venezia Giulia | 2,421 | 2.41% | 1,140 | 2.91% | 330 | 2.73% |
| Lazio | 13,268 | 2.57% | 1,742 | 2.96% | 87 | 2.80% |
| Liguria | 3,886 | 2.88% | 407 | 3.73% | 34 | 2.97% |
| Lombardy | 23,599 | 2.51% | 434 | 2.94% | 347 | 2.77% |
| Marche | 5,273 | 3.09% | 771 | 3.83% | 388 | 3.14% |
| Molise | 571 | 1.77% | 1,077 | 2.20% | 430 | 2.46% |
| Piedmont | 7,545 | 1.89% | 107 | 2.59% | 430 | 2.63% |
| Sardinia | 4,221 | 2.78% | 489 | 3.16% | 338 | 2.92% |
| Sicily | 8,280 | 1.54% | 1,197 | 1.86% | 66 | 2.21% |
| Tuscany | 9,187 | 2.65% | 1,510 | 3.01% | 359 | 2.81% |
| Umbria | 1,728 | 1.70% | 395 | 2.05% | 100 | 2.36% |
| Veneto | 8,598 | 1.84% | 2,247 | 2.28% | 131 | 2.48% |
| **TOTALE** | **111,762** | **1.92%** | **14,972** | **2.30%** | **4,171** | **2.56%** |

Finally, to compare the degree of infection transmission from students and teachers to their close contacts, we analyzed data collected by the MI from contact tracing in the monitored schools (from November 23 to December 5, 2020). Information was retrieved from 5,971 (45%) institutes in the first week and 7,035 (55.6%) institutes in the second week (public and private kindergarten, elementary and middle schools), accounting for 423,516 and 496,289 students in the first and second week, respectively. The Least Square estimates of the incidence of secondary cases over the number of tests carried out on close contacts of a positive subjects in school was less than 1% per school and week for teachers and students, in kindergarten, elementary and middle schools. Estimates of rates when the index case was a student, or a teacher were not statistically different (Table 7). The number of tests per institute per week ranged from an average of 7 in kindergarten to 18 in middle schools (Table 8). Figure 2 shows the box-plot of the numbers of test per school when a student is the index case and when the index case is a teacher. Twenty-seven schools carried out more than 100 tests in a week. Clusters, defined as >2 SARS-CoV-2 positive subjects identified in one week following contact tracing of index cases, were found in 5% to 7% of schools (Figure S2). On average, 49%-56% of all close contacts of a positive student or teacher were placed in quarantine for 10 days, with the need of a negative swab at the end of the period to be readmitted at school. These striking data corroborate the notion that elementary and middle school students not only are less infected, but also that the risk of infection at schools is low. Moreover, they indicate that contact tracing results in a high number of swabs per contacts, especially when the index case is a student, more than the average of tests for contact tracing in the general population. Altogether, our analysis of the data collected by the MI indicates that in Italy students are less infected than the general population and the overall protocols for contact tracing work, questioning whether schools played a role as amplifiers of the second Covid19 wave.

**Table 7.** Incidence of secondary infections identified during activity of contact tracing in Italian Schools (from November 23 to December 5, 2020). Incidence rates of secondary infections are defined as number of cases/number of tests occurring the same week after a SARS-COV-2 positive student or teacher was found. LSmeans, 95% Confidence Interval (95%CI) and P-values of rate of secondary infections per institute and week are estimated with multivariable generalized linear regression model. P-test indicate differences between incidence rates by type of index case.

|  | **Student as index case**<br>**LSmeans 95%CI** | **Teacher as index case**<br>**LSmeans 95%CI** | **P-test** |
| --- | --- | --- | --- |
| Kindergarten | 0.78% (0.45%, 1.20%) | 0.71% (0.33%, 1.22%) | 0.81 |
| Elementary school | 0.68% (0.48%, 0.91%) | 0.98% (0.64%, 1.39%) | 0.22 |
| Middle school | 0.74% (0.53%, 0.97%) | 0.90% (0.51%, 1.40%) | 0.50 |

**Table 8.** Activity of contact tracing following a positive case among students and teachers in Italian Schools (from 23 of November to 5 of December 2020). Mean and standard deviation of number of tests per institute

|  | Type of school | n.<br>schools | Mean number of tests | Standard deviation | Absolute<br>range |
| --- | --- | --- | --- | --- | --- |
| Student<br>Index<br>case | Kindergarten | 531 | 9 | 13 | 0-87 |
|  | Elementary | 873 | 16 | 20 | 0-150 |
|  | Middle schools | 753 | 17 | 21 | 0-87 |
| Teacher<br>Index<br>case | Kindergarten | 465 | 7 | 15 | 0-180 |
|  | Elementary | 540 | 13 | 25 | 0-232 |
|  | Middle schools | 338 | 12 | 26 | 0-117 |

**Figure 2.**
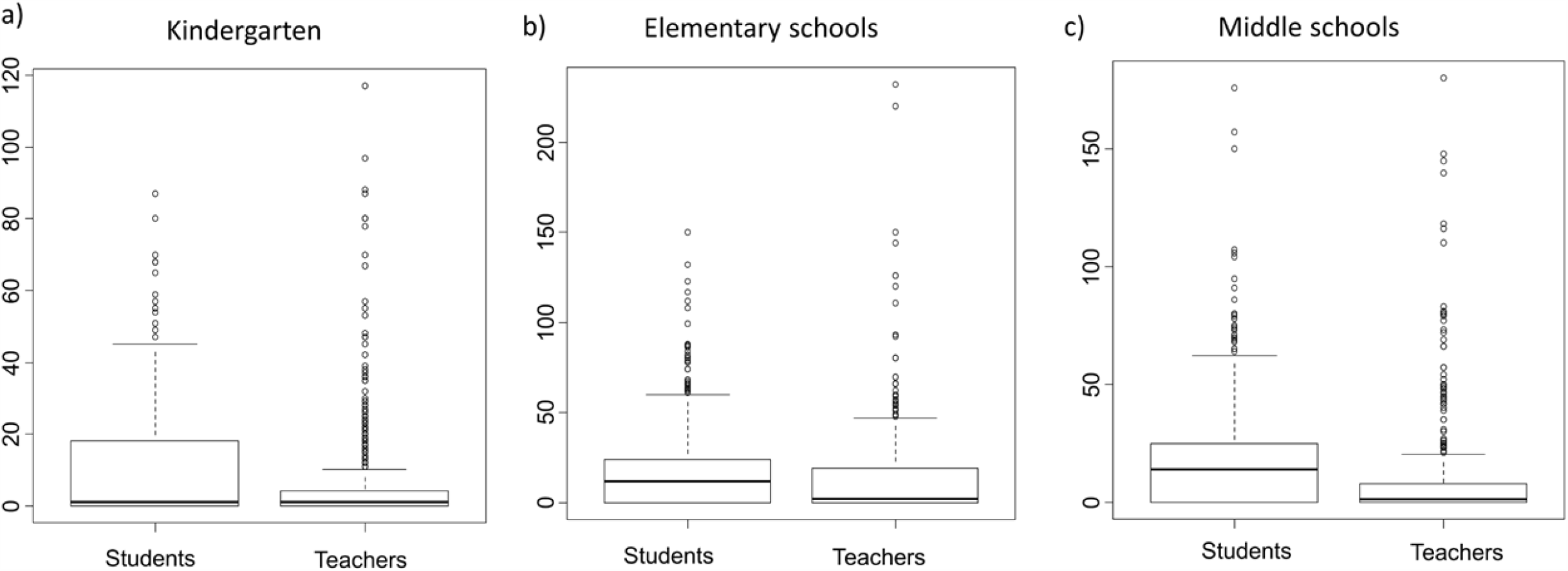
Box-plots of number of test per week per school. Boxplots of number of tests performed in the indicated type of school per week in students and teachers when they are index cases.

### Increases in R_t_ in Italian regions with different school opening dates

We reasoned that if school openings had played a role in the second wave of Covid19 in Italy, the reproduction number R_t_ shall have increased earlier in the regions where schools started earlier. We first tested this hypothesis by analyzing the case of the two provinces of Bolzano, where schools started on September 7, and Trento, where they started on September 14 (Table 2). Given the similarities between these two alpine territories in terms of orography, population density (72 inhabitants/km^2^ in Bolzano; 87 in Trento), climate and lifestyle, they represent a very useful case scenario to investigate the role of schools in the local spread of Covid19. We computed R_t_ ^21^ on the incidence of the positives at a RT-PCR for Sars-CoV-2 genetic material test from an oro/nasopharyngeal swab (data retrieved on Dec. 3, 2020 from the repository of the Italian Civil Protection Department https://github.com/pcm-dpc/Covid19). Notwithstanding that schools in Trento opened 7 days later than in Bolzano, the increase in R_t_ (defined as an increase sustained for >3 moments and leading to R_t_ >1) occurred from the period 23/9-30/9, whereas in Bolzano R_t_ started to increase from the period 29/9-6/10, suggesting that there was no temporal relation between schools opening and surge in R_t_ (Figure 3A).

**Figure 3.**
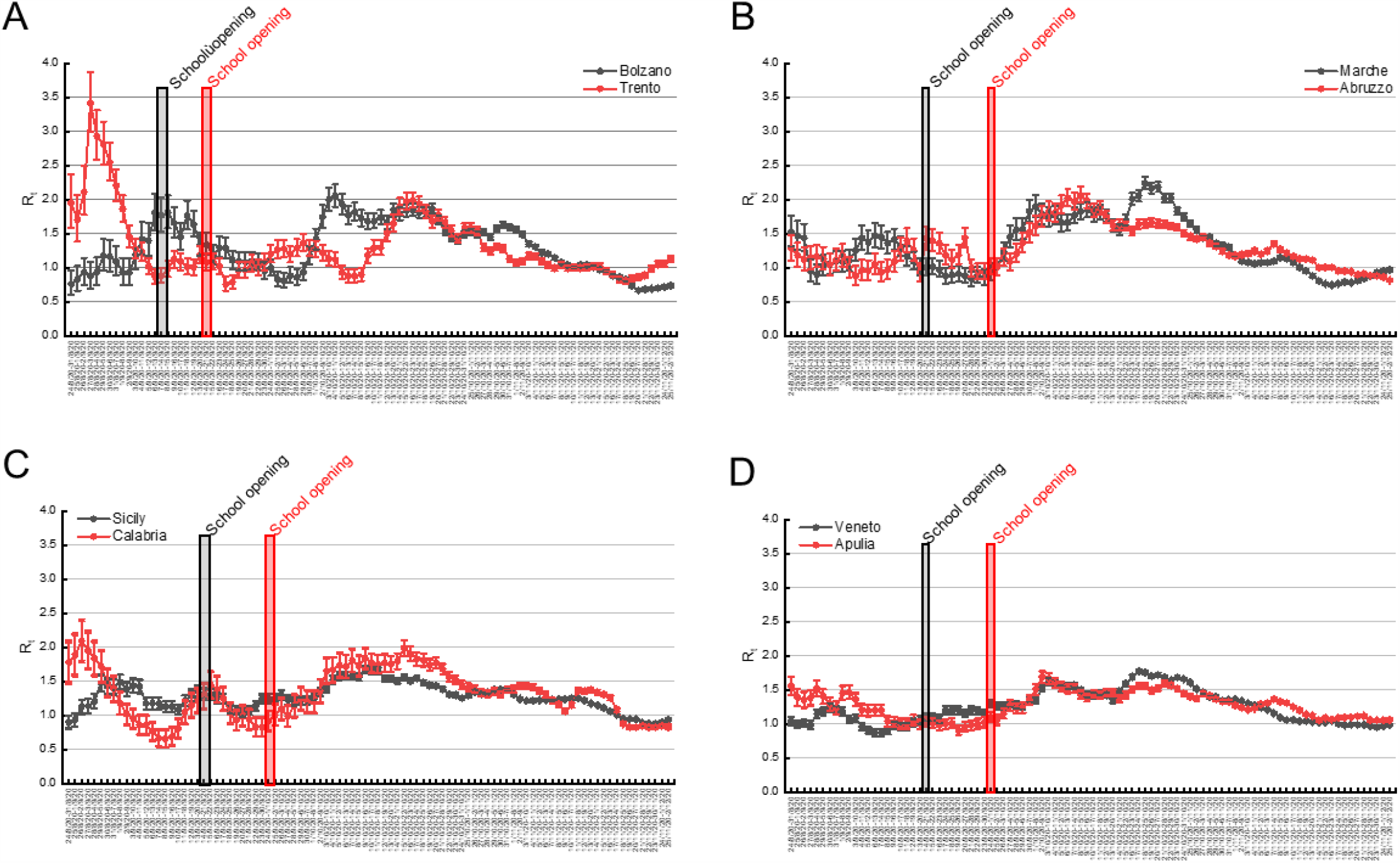
Increases in R_t_ are not univocally correlated with school opening times in different Italian territories. Pairwise comparison of median R_t_ in the indicated 7 days periods (±5-95% Confidence intervals) in the provinces of Bolzano and Trento (A) and in the indicated regions (B-D). The dates of the school openings in the respective region or province are marked with a bar of the same color. The green bars indicate the day of the entry into force of the Presidential decrees (Decree) detailed in Table 5.

We extended our analyses to different pairs of Regions where schools opened on different days. We compared the temporal distribution of R_t_ in Abruzzo and Marche, two bordering regions of central-eastern Italy. In Marche, schools opened on September 14, in Abruzzo on September 24. In both regions, R_t_ started to increase from the 25/9-2/10 period (Figure 3B). We repeated the same exercise for the pair Sicily-Calabria, where schools started on September 14 and 24, respectively. Again, we found no difference in the period when R_t_ started to increase (Figure 3C). Finally, even in the case of the pair Veneto-Apulia where schools opened on September 14 and 24 respectively, we did not appreciate any difference in the period when R_t_ started to increase (Figure 3D). Altogether, these data indicate that the increase in SARS-CoV-2 reproduction number in different Italian regions occurred indeed after school openings, but that at the same time the delay between school opening and R_t_ rise was not constant as it would be expected if it were the only driver of Covid19 diffusion. This lag time appeared indeed shorter in regions where schools opened on September 24, longer in those regions where schools opened on September 14. We further corroborated this impression by calculating the number of days from the date of the school opening to the R_t_ across all Italian regions increase (Figure S3). The average delay from school opening to R_t_ increase was 5.7 days (CI75%: 3.75-7.5) in regions where schools opened on September 22 or 24, 12.4 days (CI75%: 10-16) in regions where schools opened on September 14 or 16 (Figure 4A, P<0.05 in a Kolmogorov-Smirnov test). Conversely, the average delay between the R_t_ rise and the national election day held on September 21 was comparable in all regions: the mean was 8.7 (CI75%: 7-9) in regions where schools opened on September 22/24 and 5.2 (CI75%: 3-9) in regions where schools opened on September 7 or 14/16 (Figure 4B). In conclusion, we did not find an unequivocal delay between school opening and Rt rise.

**Figure 4.**
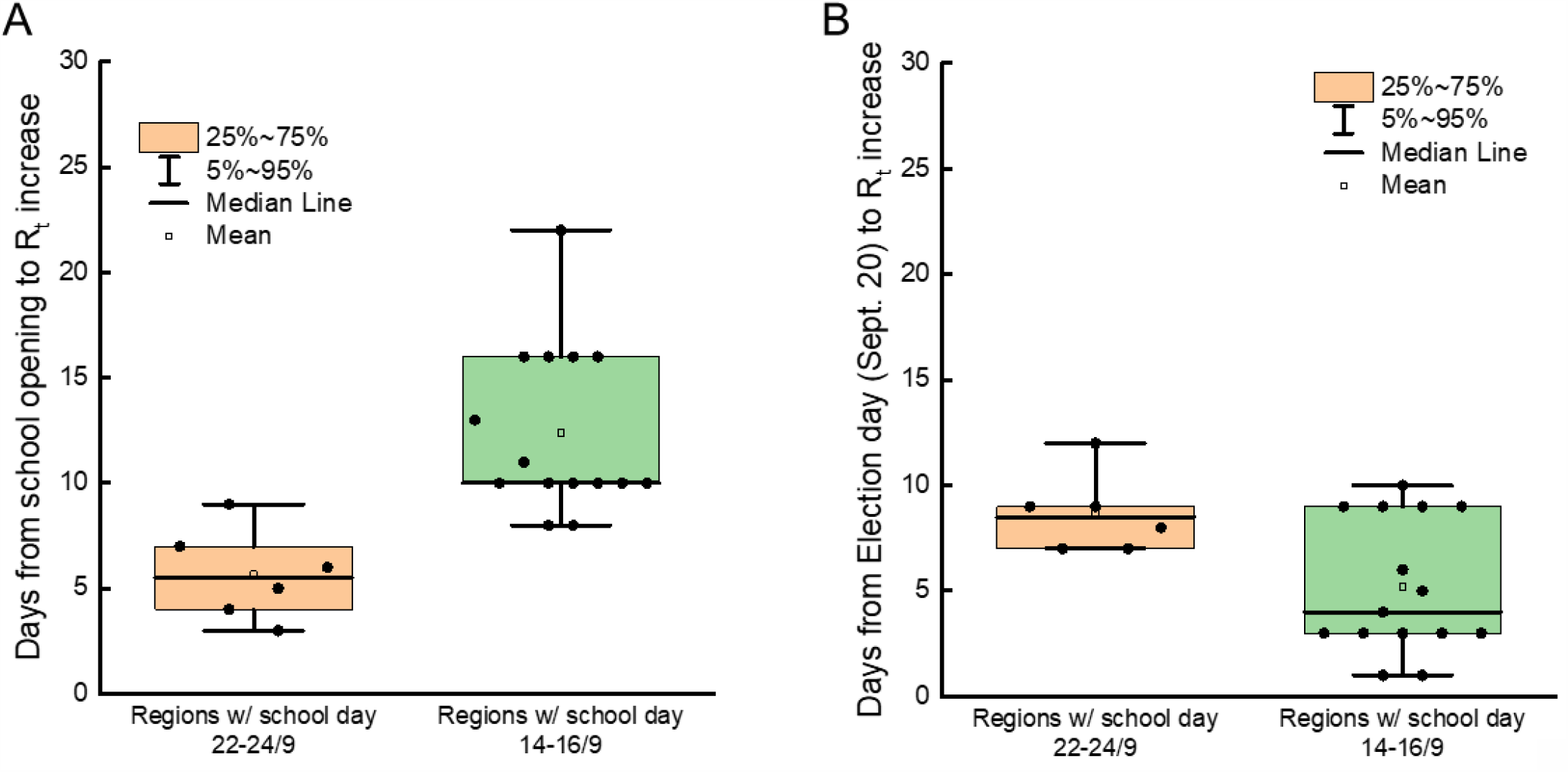
Increases in R_t_ are univocally correlated with the national election day across all Italian regions. Box plots show the indicated quantiles for the days of delay between school openings (A) and September 20 national election day (B) and R_t_ increase in Italian regions clustered by their school opening dates. Date of R_t_ increase was calculated as the first day of the period when median R_t_ started an increase sustained in time (>3 consecutives periods).

### Early increase in Covid19 incidence among adults, not school age individuals during the second wave in the Veneto region

Because we did not find a strong temporal relation between school openings and the second Covid19 wave in Italy, we decided to understand whether SARS-CoV-2 positivity circulated early in individuals different than children. To this end, we performed a prospective study on datasets extracted from the Veneto Region system of SARS-CoV-2 cases notification. We stratified incidence of newly reported Covid19 cases for age from late August to late October 2020, when overall Covid19 incidence in Veneto increased from ∼2/10,000 to ∼35/10,000. We first divided the population into infants (0-4 years of age), students (5-19), young adults (20-34), adults (35-59), seniors (60-74) and elderly (75+) and calculated daily incidence of newly reported cases in these age categories. Demographic details are reported in Table 1. Interestingly, incidence in young adults and adults increased at the end of August and then again at the end of September, whereas it remained very close to baseline in all the other age groups in these two periods (Figure 5A). This result prompted us to increase the granularity of our analysis. We stratified incidence for the classic demographic brackets (we used one single group of 75+ years old individuals as we did not find differences in incidence in groups above age 75). In late August-early September incidence increased among 45 to 49-year-old individuals and albeit less also among the 25 to 39-year-old ones. Conversely, incidence remained very low in the other analyzed age groups. Incidence increased again in the last decade of September in the age groups 45-49 and to a lower extent in the age groups 20-24 and 25-29 (Figure 5B). These data were surprising, as they suggested that at least in Veneto the earliest increase in SARS-CoV-2 positivity occurred in adults, followed by younger individuals, but not in adolescents that were often deemed as potential spreaders because of their high number of social contacts and their presumed laxity in adhering to the infection risk mitigation protocols. We therefore further inspected the temporal distribution of incidence among age classes. We generated a heatmap of the incidence of Covid19 cases in every age group in the 8 weeks under consideration, where low incidence is in green, intermediate incidence values in beige, high incidence in red. Visual inspection of this heatmap confirmed that the earliest increase in incidence occurs not among children or adolescents, but among individuals 20-49 years of age. These individuals appeared to be the drivers of the second wave, as incidence then propagated to individuals of other age categories (Figure 5C). Indeed, by applying a Euclidean distance algorithm to the same matrix used to generate the heatmap, we found that children and adolescents are ranked as the groups closest to the least affected groups by this second Covid19 wave (60-64 and 65-69 years of age). Conversely, individuals 20 to 29, and 45 to 49 years old are the most distant from the protected 60-69 years old individuals (Figure S4).

**Figure 5.**
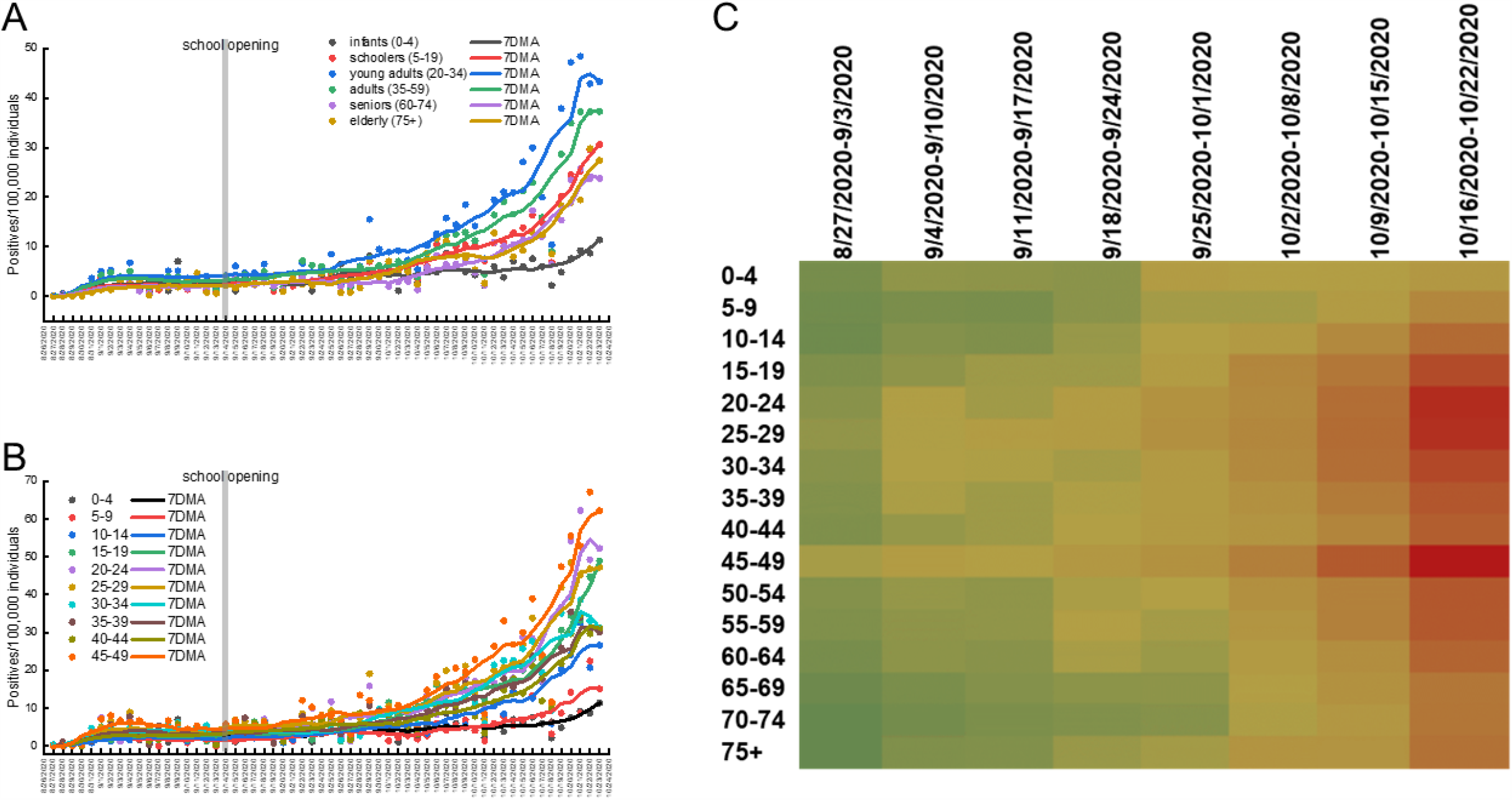
During the second Covid19 wave incidence of SARS-CoV-2 rises initially among young adults and 45-49 years old individuals in Veneto region. (A, B) Daily incidence and 7 days adjacent average (7DMA) of SARS-CoV-2 positivity among individuals of the indicated age range. Consistency of the population in each age bracket was from ISTAT and is detailed in Table 6. (C) Heatmap of weekly incidence of SarsCoV2 in individuals of the indicated age ranges in the Veneto region during the indicated timeframe. The color scale goes from green (low incidence) to beige (medium incidence) and to red (high incidence).

Finally, we compared the incidence of SARS-CoV-2 from September 19 to the October 18 among teachers and among the general population of the age group 25-65 in Veneto. We selected this age group because teachers’ age is comprised between these two extremes, given the required tertiary education to be enrolled, and the legal retirement age of teachers. Interestingly, incidence among teachers starts to increase after the general population of the same age; moreover, at the end of the period under consideration, incidence among teachers and among the general population aged 25-65 is not significantly different (12/10,000 vs. 11.1/10,000, P=0.36, Figure S5). We also investigated the frequency of secondary infections at schools in Verona and province from November 25 to December 21 on datasets extracted from the Veneto Region system of SARS-CoV-2 cases notification. We found 380 student, 30 staff and 114 teachers index cases in 339 schools for which contact tracing was performed. From this contact tracing and testing, a total of 76 secondary cases were identified (Table 9). The frequency of secondary cases was higher among students than among teachers and staff (71%, 22.4% and 6.6%, respectively). A secondary case among teachers was more frequent when the index case was a teacher than when it was a student (38% vs. 11%, P=0.007). Secondary cases among staff members were exclusively due to contacts with other staff members.

**Table 9.** Index and secondary cases in 339 schools of the Province of Verona (from November 25 to December 21, 2020). Note that in the case of one teacher index case, 2 secondary cases among other teachers were identified. Frequency of teachers and students is significantly different by index case: P=0.007 students vs teachers.

| Index cases |  | Secondary cases |  |  |  |
| --- | --- | --- | --- | --- | --- |
|  |  | Total | Students | Teachers | Staff |
| Students | 355 | 60<br>(100%) | 54<br>(90%) | 6<br>(10%) | 0<br>(0%) |
| Students age <13 |  | 38<br>(100%) | 33<br>(87%) | 5<br>(13%) | 0<br>(0%) |
| Students age 13-18 |  | 22<br>(100%) | 21<br>(95%) | 1<br>(5%) | 0<br>(0%) |
| Teachers | 112 | 16<br>(100%) | 10<br>(63%) | 6<br>(37%) | 0<br>(0%) |
| Staff | 25 | 5<br>(100%) | 0<br>(0%) | 0<br>(0%) | 5<br>(100%) |
| Total | 492 | 81 | 64 | 12 | 5 |

Altogether, these analyses indicate that in the Italian Veneto region, children and adolescents were not early drivers of the second wave, which was conversely associated with an early increase in incidence among 20-29- and 45-49-years old individuals. Furthermore, teachers were not at greater risk than the age matched general population and when teachers were infected at school, infections were due mainly to other teachers.

### School closures did not alter the rate of R_t_ decline in Lombardy and Campania

Because we did not find a correlation between school opening and the rise in R_t_, we wished to understand whether the opposite, i.e., school closures, impacted on R_t_. Again, the territorial differences in the mandate of different NPI in Italy offers a useful paradigm to investigate this possibility. We considered here the two cases of Lombardy, where the Governor mandated closure of high schools from October 26; and Campania, where the Governor mandated the closure of all school grades (including kindergartens) from October 16. Lombardy and Campania together account for 25% of Italy’s population, being the first and second most populous regions. These school closures occurred before the national Government implemented a regional risk stratification system to modulate lockdown according to the local epidemiological and hospital stress status (November 6), but after the mandate for universal mask wearing outside of home (October 14) and in the case of Lombardia after the closure of restaurants, cafes, and bars at 6PM with a nationwide curfew at 10PM (October 23). Because of the stability (i.e., lack of recalculations) of the data communicated by these two regions, we could compute Rt on the positives at a RT-PCR for Sars-CoV-2 in swabs prescribed by a physician (*sospetto diagnostico*, i.e. clinical indication). Interestingly, R_t_ decline started well before high school closures in both regions: in Lombardy in the October 8-15 period (Figure 6A for absolute R_t_ values and 5B for its first order derivative); in Campania, in the September 30-October 7 period (Figure 6C for absolute R_t_ values and 5D for its first order derivative). Noteworthy, the same pattern was observed if we analyzed R_t_ computed over total Sars-CoV-2 positivity, albeit in the case of Campania R_t_ decline started only three moments before the implementation of school closures (Fig. S3, red lines in the plots of Campania and Lombardy). In the case of Campania, we could also extend our analysis to the overall incidence among students and general population. We found that while incidence dropped among students, because they were no longer attending schools and therefore tested, the overall incidence in the general population continued to increase (Figure S6), reflecting the fact that R_t_ remained >1 until the 5-11 November period. Altogether, these data indicate that school closures did not impact on the speed of R_t_ decline in Lombardy and Campania. Furthermore, the increasing trend of Covid19 incidence in the general population observed in Campania was not curtailed by school closures.

**Figure 6.**
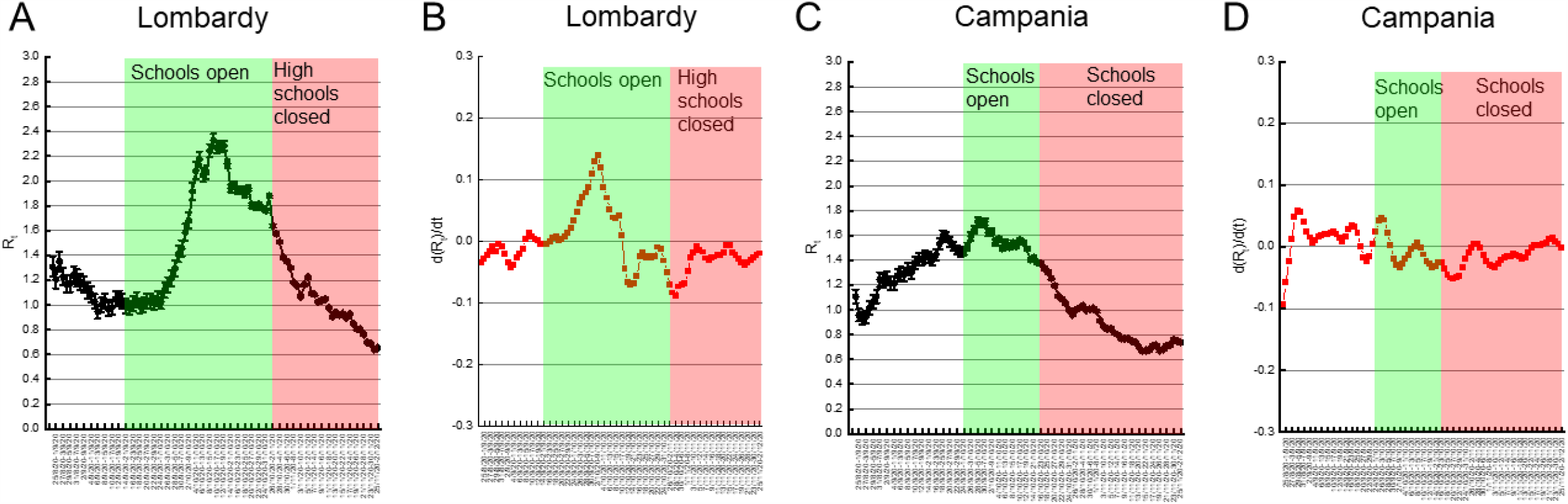
School closures do not affect R_t_ decrease in Lombardy and Campania. (A, C) Median R_t_ in the indicated 7 days periods (±5-95% Confidence intervals) in Lombardy (A) and Campania (C). Days of school opening and closure are indicated. (B, D) First order derivative of R_t_ in Lombardy (B) and Campania (D). Days of school opening and closure are indicated.

## Discussion

Whether school reopening contributed to the second wave of Covid19 in Italy was unclear. Here, by analyzing data from Italian regions and schools, we did not find a significant association between school opening and rise of infection in the general population. Our conclusion is based (i) on the finding of lower incidence of SARS-CoV-2 positivity among students than in the general population; (ii) on the lack of association between staggered school reopening in different Italian regions and direction change in reproduction number R_t_; and (iii) on the analysis of the temporal changes in the incidence among different age classes in the Veneto region between late August and late October 2020.

At variance with influenza, where younger individuals seem to represent a reservoir of virus and contribute to its propagation to the general population ^23-27^ SARS-CoV-2 seems to spare school age children and adolescents: clinically, they are mostly paucisymptomatic ^5^; from the epidemiology of infection perspective, they are very rarely accounted for as the index case ^11^, indicating that not only they are largely spared from the clinical consequences of the infection, but they are also less likely to transmit it. Overall, these data suggest that spread of Covid19 within school settings may be limited ^28,29^. Indeed, our data indicate that infection incidence is lower in students of any cycles, compared to the general population. Moreover, at least in the case of elementary school children, contact tracing in schools confirms that they are less likely to transmit the virus, as evidenced by a 27% reduction in the number of secondary cases among teachers when the index case is a student (10%) compared to the number of secondary cases elicited when the index case is a student (37%). These epidemiological data are in line with the finding that children harbor antibodies against the common other betacoronaviruses, and that these antibodies are cross reactive and neutralizing against SARS-CoV-2 ^7^. Our findings are also consistent with several other reports of very limited spread of Covid19 between children and from children to adults. In

Australia (New South Wales), following Covid19 positivity of 9 students in primary and high schools and 9 staff members, only 2 of the 735 students, and 0 of the 128 staff members with whom they had contact were identified as secondary cases. In Ireland, during the first wave, 6 Covid19 cases were identified in schools (three children and three adults). Among their 1,155 school contacts, zero infections were recorded ^30^. In the Netherlands, ten Covid19 cases aged <18 had 43 contacts, but nobody was infected, whereas 221 patients older than 18 were associated with 8.3% of infections ^31^.

We found higher rates of incidence in teachers and school staff compared to the general population. One possible explanation for this finding is that school environments are meticulously and continuously controlled, confirmed by the number of tests performed for each positive case. Indeed, in Campania where schools were open for a mere 17 school days (from September 24 to October 16; school week of 5 days), incidence among teachers and school staff in the September 12-November 7 period is still higher than that in the general population. It would be difficult to ascribe this difference to 17 days of school over a total of 56 days. It cannot be argued that teachers and staff members are more susceptible to infection than the general population; in fact, this increase in the incidence of test positives is not mirrored by an increase in mortality-morbidity that would mark a more susceptible population ^32^. Another possibility is that teachers become infected at school. Judging from contact tracing activity in schools of the populous province of Verona (Veneto), this occurrence is however quite rare: only 13 teachers were accounted for as secondary cases from 524 traced index cases.

Decision makers, popular press and public opinion in Italy ascribe the second wave of Covid19 to school reopening. This is often accompanied by deprecating comments on “individual behavior” of adolescents especially, who would not follow the strict rules at school or outside them. However, our data suggest that this common sentiment is not evidence- based, but perhaps grounded on the temporal correlation between school opening (in September) and second wave (in October-November). Indeed, our data do not identify an association between school reopening and rise in R_t_ analyzed on a regional basis. Because of the staggered school reopening calendar in Italy, we were well positioned to address whether there was an association between the date of school opening and the date of reproduction number increase. We did not find such association that was conversely present when we analyzed the temporal distance between R_t_ rise and the election day held in Italy on September 20 (and morning of 21), 2020.

Other reports from across the world are in line with our findings: in Great Britain, staff had higher incidence than students (27 cases [95% CI 23–32] per 100,000 per day among staff compared with 18 cases [14–24] in early years students, 6.0 cases [4.3–8.2] in primary schools students, and 6.8 cases [2.7–14] in secondary school students]) and most cases linked to outbreaks were in staff members (154 [73%] staff vs 56 [27%] children of 210 total cases). The median number of secondary cases in outbreaks was one (IQR 1–2) for student index cases and one (1–5) for staff index cases ^33^. In Spain as well, the evolution of the global incidence does not suggest significant effects of the reopening of schools. In most cases there was no or a slight increase in pediatric cases, consistent with the diagnostic efforts in schools ^34^. In Germany, data collected from 53,000 schools and daycares in autumn indicated that only an average of 32 schools one week had more than two positives ^35^

A different question is whether closing schools is efficacious in curtailing viral spread. In some Italian regions analyzed here, school closure was mandated by local authorities and finally in certain regions by the National Government. However, this closure had no effect on the incidence of Covid19 in the general population or in the decline of R_t_, which had started before the mandated school closure and that continued with the same speed, irrespective of school closures in Lombardy (partial) and Campania (total). This finding is in line with a literature review of all available studies (n=16) on the efficacy of school closures and other social distancing practices in schools in China and Hong Kong, where the rapidly implemented school closures did not substantially contribute to the control of the spread ^15^. In Australia, by comparing data from 25 schools of different grades with those of the general population, it was found that students and school staff did not contribute to the spread of the virus more than the general population ^36^. On the other hand, an analysis of the impact of different NPI on the reproduction number R_t_ across 131 countries found that at day 28 after the implementation, school closures alone could reduce R_t_ by 15% (R ratio: 0.85, 95%CI: 0.66-1.10), whereas school reopening could increase it by 24% (R ratio: 1.24, 95%CI: 1.00- 1.52). However, these estimates are affected by a very large confidence interval, hence are not statistically significant. Moreover, authors warn on the limitations of their estimates: for example, they could not consider the different precautions related to the reopening of schools taken by some countries, such as physical distancing within classrooms and masking procedures; they did not consider the impact of school holidays and the effect of reopening different school levels (e.g., elementary and middle schools). Finally, authors analyze the impact of given NPIs by comparing R_t_ from two arbitrarily drawn periods before and after the implementation of the given NPI. While this approach might be more practical when comparing multiple countries, it is less informative than our analysis performed overall the R_t_ curve. Interestingly, we found that R_t_ started declining even before the implementation of any NPI, in all the regions analyzed. In certain cases, like the Province of Trento, the nationally implemented NPI entered in force after R_t_ had declined below the threshold of 1. These results, while perhaps surprising, are in line with findings from the group of Merler, who analyzed the impact of the national March-May lockdown on R_t_ in Italy. While they concluded that lockdown reduced R_t_ and brought it below 1, they admitted that the decline in R_t_ had started well before the national lockdown was implemented. Visual inspection of their published R_t_ curves indeed confirms that this extreme NPI did not affect the slope of R_t_ decline ^37^.

Our study is strengthened by the several sources of data used. Longitudinal data of regional incidence of SARS-COV-2 positives subjects deposited in the public repository of the Italian Civil Protection, incidence from the Veneto Region system of Covid19 case notification with information by age, and incidence in schools from MI with information for students, teachers and non-teaching staff.

Our study also suffers from some limitations. Information on SARS-CoV-2 positive individuals in schools are retrieved by school Principals. Second, these data represent the situation of the whole school, not of the single subjects. Third, data on number of SARS-COV-2 positives subjects deposited in the public repository of the Italian Civil Protection might suffer from delays in reporting or even worse from differences in reporting criteria by different regions. It shall be however noted that the nationwide R_t_ computed on the total positives and that on the cases by diagnostic suspicion are very similar and that their temporal trends are superimposable, thus reinforcing the strength of the analysis presented here.

In conclusion, our analysis does not find an association in Italy between dates of school opening and the increase in SARS-CoV-2 R_t_. Reciprocally, school closures did not affect the rate of R_t_ decline. Also, the incidence of SARS-CoV-2 among students is lower than that in the general population; In addition, the incidence among teachers is greater than the incidence in the general population, but comparable to that of recorded in the general population of the same age. Finally, contact tracing in schools resulted in very low frequency of secondary infections found per test, and low frequency of clusters despite a high number of tests every week. Our analysis provides evidence that school openings are not to be considered as a relevant factor influencing the spread of the Covid19 epidemics and that school closures did not improve the already occurring decline in the reproduction number of Covid19, at least in two populous Italian regions. Closure of schools has dire consequences on children and adolescents motor activity, social interaction, psychological well-being ^38,39^ and psychopathological problems ^40,41^, on the risk of obesity ^42^ and screen addiction ^43^, on the protection from situations of domestic abuse ^44^, and on learning performance. Our data add further support to the consolidating notion that risks of school closures are not outweighed by benefits. They moreover suggest that the conclusion that school openings favored Covid19 spread is correlative at best and hence it does not help in the identification of the best NPIs to curtail SARS-CoV-2 diffusion.

## Data Availability

Data are retrieved from public access databases

https://github.com/pcm-dpc/COVID-19

## Acknowledgments

This work was partially supported by the Italian Ministry of Health with Ricerca Corrente and 5×1000 funds (to SG). Federica Bellerba is a PhD student at the European School of Molecular Medicine (SEMM), Milan, Italy. We thank Dr. Guido Silvestri (Emory University School of Medicine) for his support.

