## Supplementary Figures and tables for "No evidence of association between schools and SARS-CoV-2 second wave in Italy"

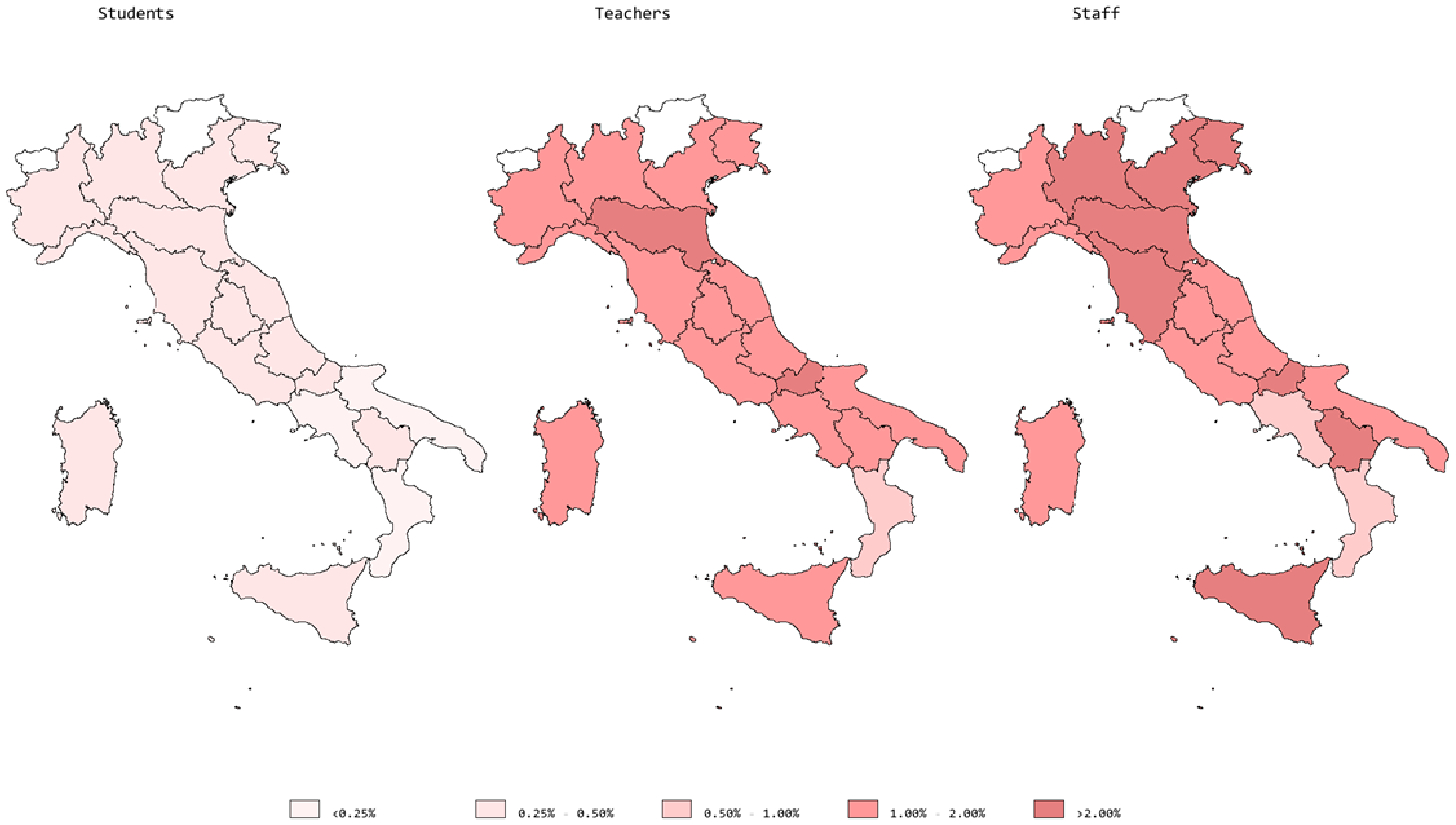


**Figure S1. Maps of incidence of SARS-Cov-2 positive students, teachers and other staff in Italian regions.** Incidence of SARS-CoV-2 positive cases in elementary and middle schools in the week 23-28 November, 2020 from the Ministry of Education database. No data is available for Valle d'Aosta, Trento and Bolzano.

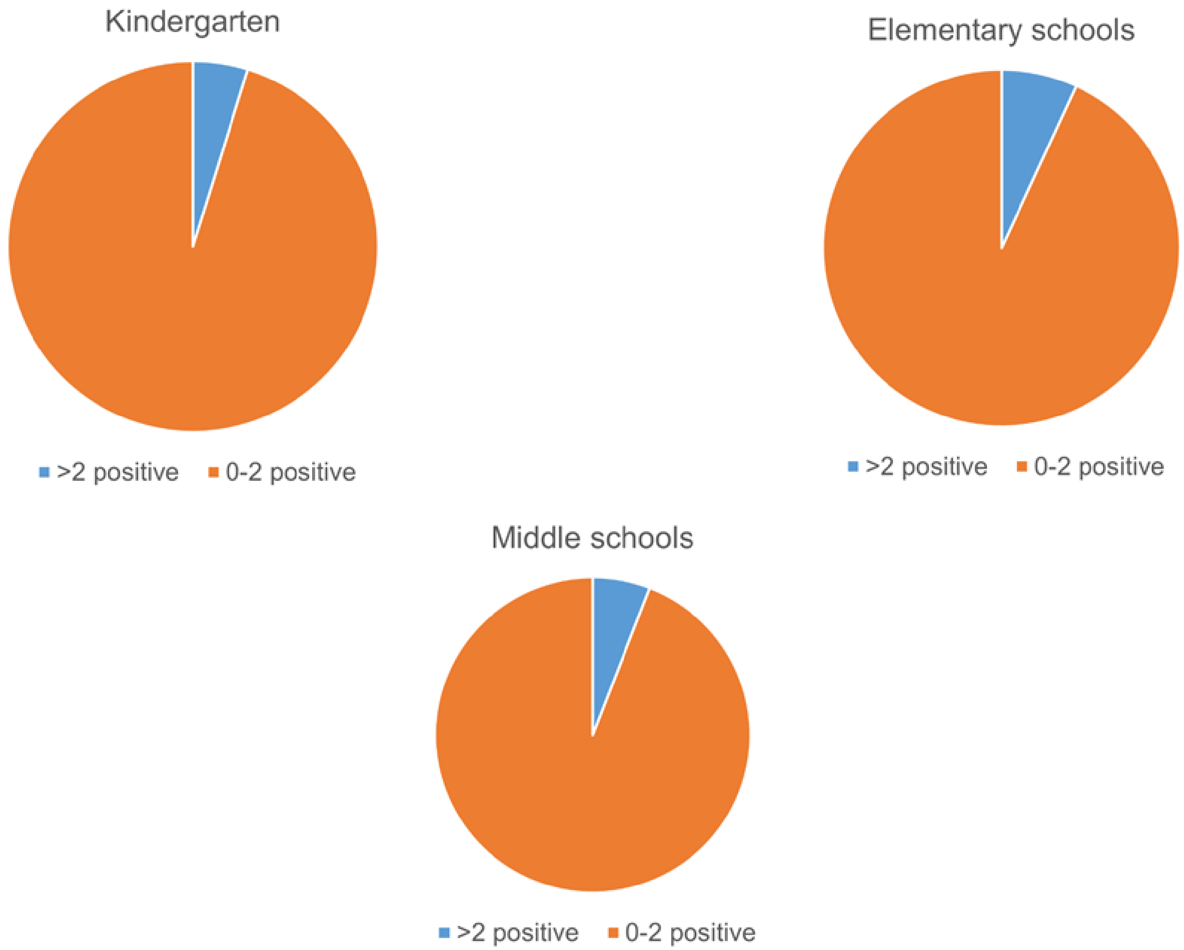


Figure S2. **Pie charts of occurrence of clusters (>2 secondary cases) in schools.** The percentage of schools where more than 2 Sars-Cov-2 positive cases identified among close contacts of a school index case is indicated.

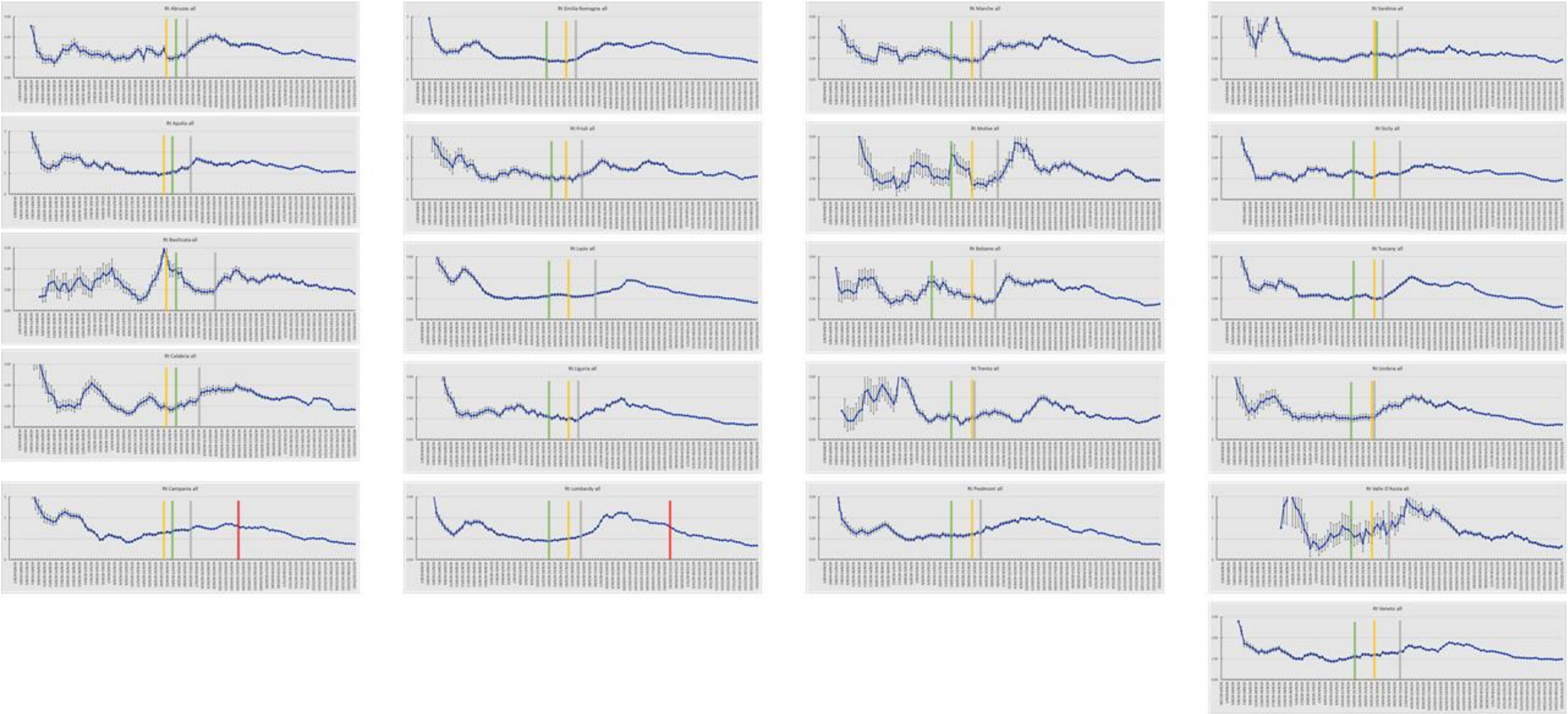

Figure S3. **$R_t$ curves across all Italian regions and autonomous provinces.** The green lines indicate school opening in the individual region; the yellow lines the national election day; they grey lines the moment when starts to increase, defined as a sustained increase for >3 consecutive moments. For Lombardy and Campania, the red lines indicate dates of school closures implemented by the local authorities.

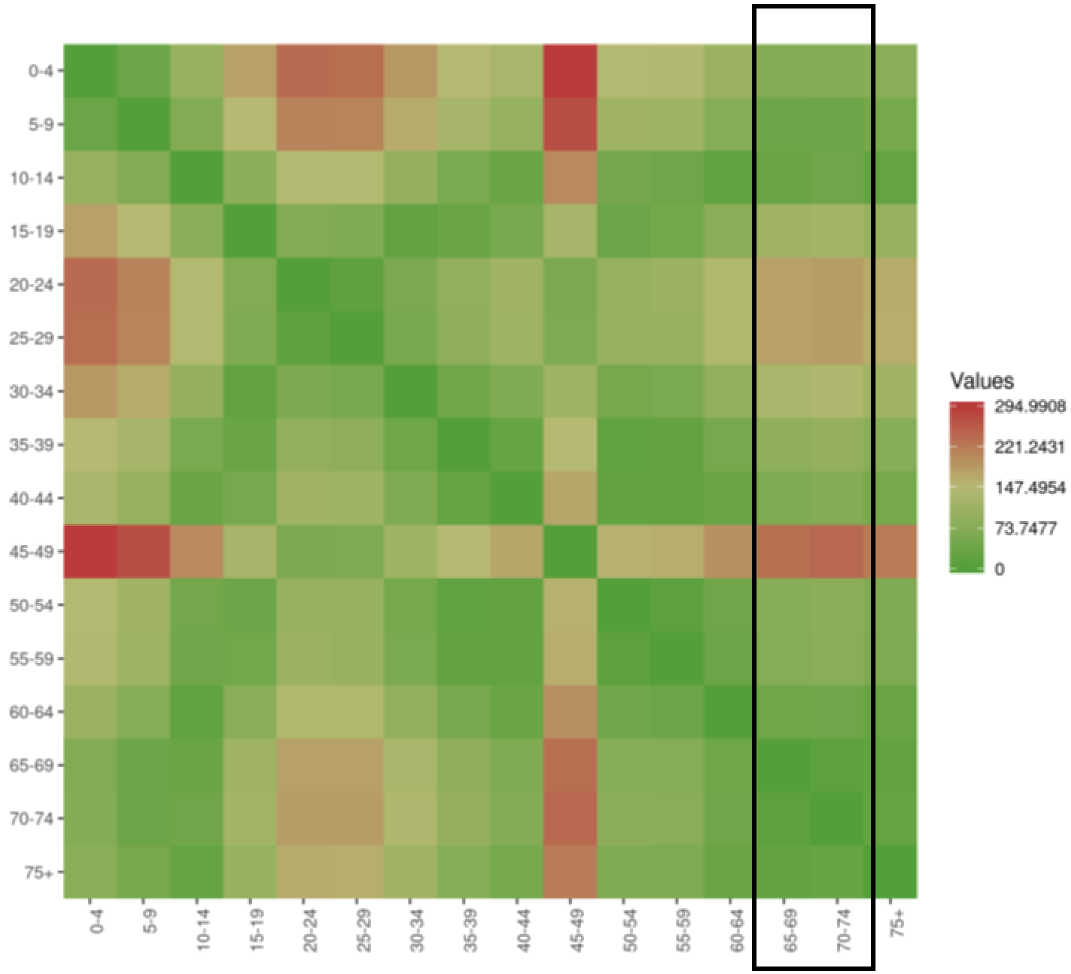


Figure S4. **Heatmap of the Euclidean distance algorithm of incidence of Covid-19 by age.** Data are from Fig. 5. The box highlights the distance between the different age groups and the groups of age 65-74 that are the ones where prevalence is the lowest.

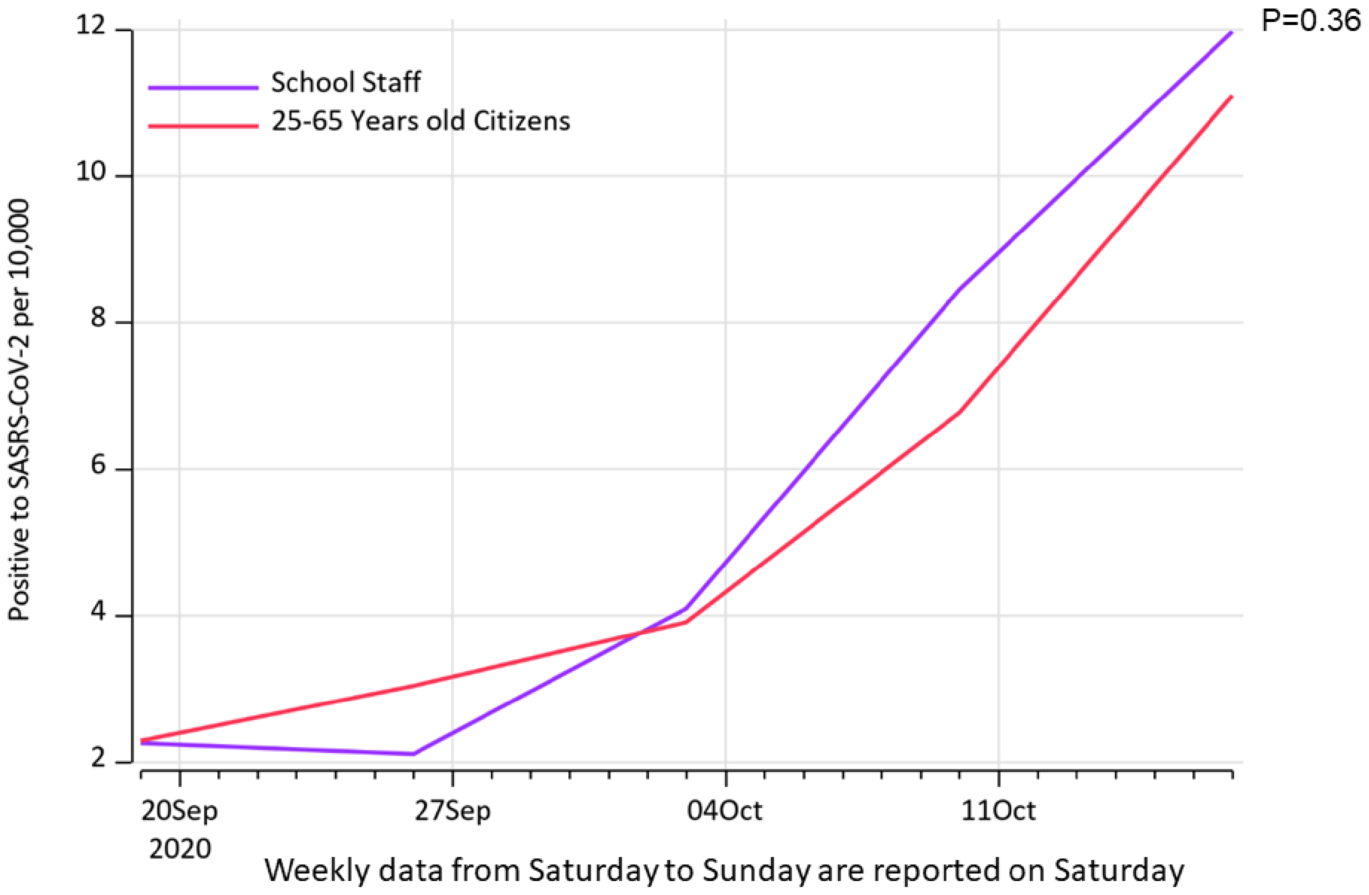


Figure S5: **Incidence of SARS-Cov-2 among teachers and the general population of the age interval 25-65 in Veneto.**

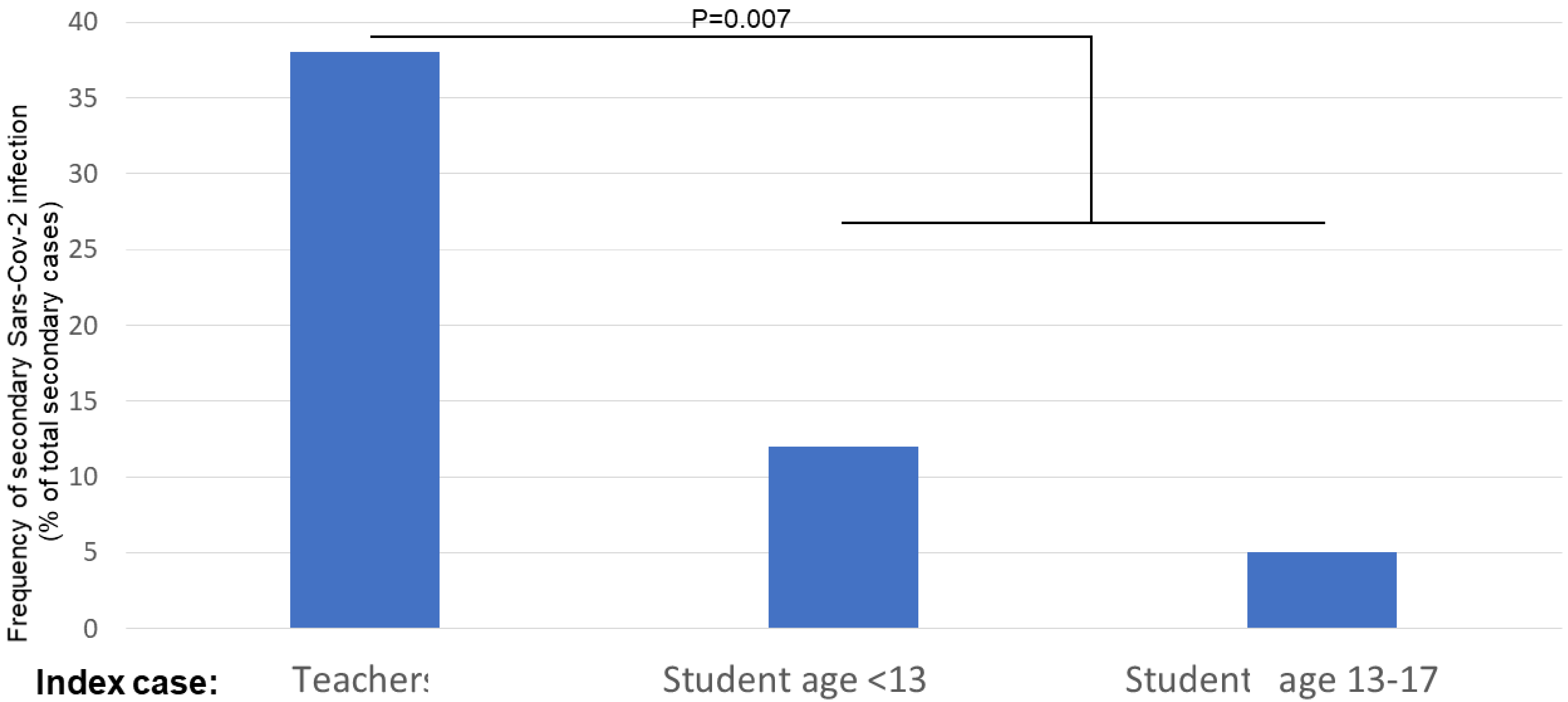


Figure S6. **Teachers are more often positive to Sars-Cov-2 if the index case at school is a teacher.** P-value from a Chi-square test for type of index case: teacher vs student.

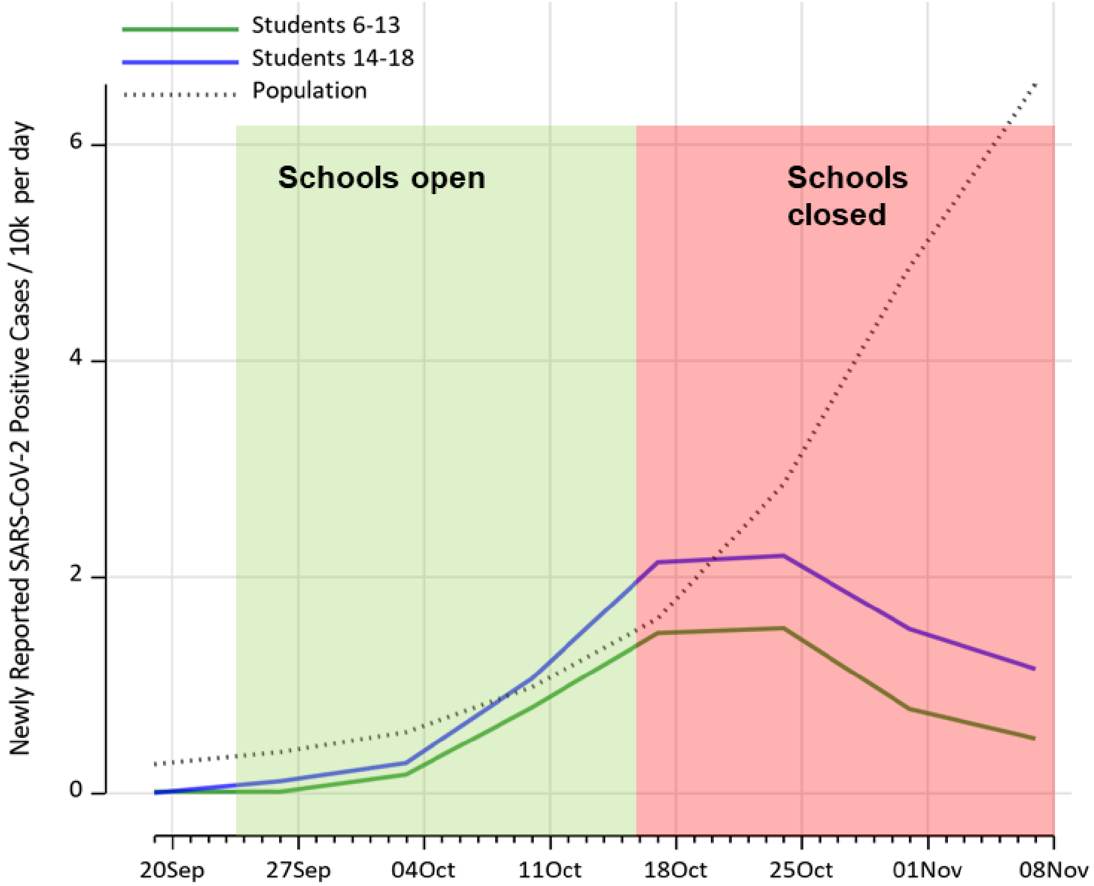


**Figure S7. Closure of schools in Campania did not curtail the diffusion of Sars-CoV-2 among the general population.** Incidence in Campania of Sars-CoV-2 among students of the indicated age (6-13: primary and middle schools; 14-18: high schools) and in the general population of the region. Boxes highlight times of school opening and closure. School closure resulted in a drop in cases among students but not among the general population.

**Supplementary Tables**

| REGION | New positive among students | % of positive students | New positive among teachers | % of positive teachers | New positive among other personnel | % of positive people among other personnel |
|---|---|---|---|---|---|---|
| **Basilicata** | 9 | 0.98% | 3 | 3.53% | 4 | 5.71% |
| **Umbria** | 20 | 0.89% | 10 | 3.48% | 21 | 2.42% |
| **Abruzzo** | 18 | 0.47% | 30 | 2.26% | 59 | 2.41% |
| **Lazio** | 170 | 0.46% | 370 | 2.06% | 7 | 2.31% |
| **Emilia Romagna** | 178 | 0.44% | 146 | 1.96% | 6 | 2.31% |
| **Veneto** | 255 | 0.34% | 83 | 1.89% | 46 | 2.06% |
| **Toscana** | 61 | 0.32% | 1 | 1.56% | 15 | 1.94% |
| **Friuli Venezia Giulia** | 41 | 0.32% | 74 | 1.53% | 79 | 1.85% |
| **Lombardia** | 505 | 0.32% | 14 | 1.53% | 146 | 1.83% |
| **Puglia** | 35 | 0.25% | 23 | 1.51% | 1 | 1.79% |
| **Piemonte** | 94 | 0.23% | 34 | 1.35% | 21 | 1.71% |
| **Sardegna** | 17 | 0.21% | 8 | 1.26% | 10 | 1.69% |
| **Sicilia** | 42 | 0.20% | 54 | 1.21% | 30 | 1.26% |
| **Marche** | 5 | 0.16% | 29 | 0.93% | 19 | 1.08% |
| **Liguria** | 13 | 0.15% | 7 | 0.75% | 27 | 0.92% |
| **Calabria** | 4 | 0.07% | 5 | 0.69% | 6 | 0.84% |
| **Campania** | 8 | 0.02% | 6 | 0.55% | 2 | 0.30% |
| **Molise** | 0 | 0.00% | 16 | 0.29% | 0 | 0.00% |
| **TOTAL** | **1,475** | **0.30%** | **913** | **1.58%** | **499** | **1.67%** |

Table S1. Incidence in students, teachers and other personnel by region, from kindergarten to middle school (private schools)

| REGION | Cases of quarantine among students | % of quarantine among students | Cases of quarantine among teachers | % of quarantine among teachers | Cases of quarantine among other personnel | % of quarantine among other personnel |
|---|---|---|---|---|---|---|
| **Marche** | 183 | 5.78% | 5 | 5.88% | 5 | 7.14% |
| **Friuli Venezia Giulia** | 525 | 4.06% | 84 | 5.52% | 57 | 6.57% |
| **Sardegna** | 319 | 4.02% | 73 | 5.51% | 33 | 4.60% |
| **Basilicata** | 34 | 3.68% | 14 | 4.88% | 83 | 3.72% |
| **Veneto** | 2,564 | 3.40% | 44 | 4.81% | 71 | 2.98% |
| **Emilia Romagna** | 1,377 | 3.38% | 209 | 4.33% | 121 | 2.83% |
| **Toscana** | 628 | 3.30% | 275 | 3.69% | 21 | 2.71% |
| **Puglia** | 461 | 3.29% | 23 | 3.63% | 7 | 2.69% |
| **Lazio** | 1,057 | 2.89% | 524 | 2.92% | 6 | 1.98% |
| **Lombardia** | 4,349 | 2.72% | 126 | 2.87% | 157 | 1.96% |
| **Piemonte** | 1,027 | 2.54% | 63 | 2.51% | 23 | 1.88% |
| **Abruzzo** | 87 | 2.27% | 13 | 1.79% | 1 | 1.79% |
| **Umbria** | 46 | 2.05% | 76 | 1.70% | 35 | 1.19% |
| **Sicilia** | 336 | 1.56% | 1 | 1.56% | 7 | 1.18% |
| **Liguria** | 120 | 1.42% | 12 | 1.28% | 20 | 1.14% |
| **Molise** | 1 | 0.15% | 21 | 0.67% | 6 | 0.89% |
| **Calabria** | 3 | 0.05% | 7 | 0.64% | 21 | 0.86% |
| **Campania** | 12 | 0.03% | 19 | 0.34% | 1 | 0.46% |
| **TOTAL** | **13,129** | **2.65%** | **1,589** | **2.74%** | **675** | **2.27%** |

Table S2 Quarantines in students, teachers and other personnel by region, from kindergarten to middle school (private schools)

| REGION | Kindergarten | | | | Elementary schools | | | |
|---|---|---|---|---|---|---|---|---|
| | New positive among students | % of positive students | New positive among teachers | % of positive teachers | New positive among students | % of positive students | New positive among teachers | % of positive teachers |
| **Abruzzo** | 6 | 0.29% | 7 | 3.04% | 12 | 1.06% | 1 | 0.64% |
| **Basilicata** | 9 | 1.10% | 3 | 4.11% | 0 | 0.00% | 0 | 0.00% |
| **Calabria** | 3 | 0.08% | 5 | 1.07% | 1 | 0.12% | 1 | 0.75% |
| **Campania** | 5 | 0.03% | 11 | 0.68% | 3 | 0.02% | 4 | 0.25% |
| **Emilia Romagna** | 79 | 0.33% | 46 | 1.69% | 49 | 0.58% | 11 | 1.15% |
| **Friuli Venezia Giulia** | 23 | 0.25% | 25 | 2.84% | 6 | 0.32% | 1 | 0.47% |
| **Lazio** | 17 | 0.19% | 15 | 1.61% | 73 | 0.51% | 19 | 1.23% |
| **Liguria** | 5 | 0.11% | 2 | 0.49% | 6 | 0.28% | 3 | 1.18% |
| **Lombardia** | 210 | 0.25% | 210 | 2.75% | 107 | 0.36% | 66 | 1.85% |
| **Marche** | 3 | 0.18% | 4 | 2.58% | 1 | 0.22% | 1 | 1.23% |
| **Molise** | 0 | 0.00% | 1 | 1.75% | 0 | 0.00% | 0 | 0.00% |
| **Piemonte** | 27 | 0.13% | 50 | 2.54% | 24 | 0.27% | 22 | 2.16% |
| **Puglia** | 23 | 0.23% | 18 | 2.20% | 10 | 0.31% | 5 | 1.20% |
| **Sardegna** | 10 | 0.19% | 13 | 2.60% | 4 | 0.25% | 1 | 0.60% |
| **Sicilia** | 13 | 0.11% | 7 | 0.76% | 20 | 0.37% | 15 | 2.78% |
| **Toscana** | 13 | 0.13% | 14 | 1.28% | 38 | 0.57% | 14 | 1.76% |
| **Umbria** | 9 | 0.55% | 2 | 1.06% | 11 | 2.89% | 8 | 14.29% |
| **Veneto** | 131 | 0.24% | 100 | 2.21% | 63 | 0.64% | 21 | 1.94% |
| **TOTAL** | **586** | **0.22%** | **533** | **2.12%** | **428** | **0.39%** | **193** | **1.53%** |

Table S3 Incidence in students and teachers by region in kindergarten and elementary school (private schools)

| | Middle school | | | |
|---|---|---|---|---|
| **REGION** | **New positive cases among students** | **% of positive students** | **New positive cases among teachers** | **% of positive teachers** |
| **Abruzzo** | 110 | 0.43% | 70 | 2.17% |
| **Basilicata** | 117 | 1.03% | 31 | 2.09% |
| **Calabria** | 81 | 0.18% | 64 | 1.08% |
| **Campania** | 188 | 0.13% | 168 | 1.00% |
| **Emilia Romagna** | 679 | 0.70% | 160 | 1.83% |
| **Friuli Venezia Giulia** | 123 | 0.52% | 39 | 1.67% |
| **Lazio** | 642 | 0.54% | 150 | 1.13% |
| **Liguria** | 224 | 0.70% | 48 | 1.43% |
| **Lombardia** | 1,014 | 0.44% | 454 | 1.92% |
| **Marche** | 183 | 0.47% | 64 | 1.64% |
| **Molise** | 55 | 0.78% | 17 | 1.99% |
| **Piemonte** | 295 | 0.32% | 309 | 2.98% |
| **Puglia** | 175 | 0.17% | 136 | 1.22% |
| **Sardegna** | 157 | 0.46% | 65 | 1.46% |
| **Sicilia** | 715 | 0.58% | 212 | 1.36% |
| **Toscana** | 327 | 0.44% | 166 | 2.07% |
| **Umbria** | 59 | 0.27% | 20 | 0.85% |
| **Veneto** | 775 | 0.71% | 179 | 1.60% |
| **TOTAL** | **5,919** | **0.45%** | **2,352** | **1.60%** |

Table S4 Incidence in students and teachers by region in middle school (private schools)
